# Multimodal BEHRT: Transformers for Multimodal Electronic Health Records to predict breast cancer prognosis

**DOI:** 10.1101/2024.09.18.24312984

**Authors:** Ndèye Maguette Mbaye, Michael Danziger, Aullène Toussaint, Elise Dumas, Julien Guerin, Anne-Sophie Hamy-Petit, Fabien Reyal, Michal Rosen-Zvi, Chloé-Agathe Azencott

## Abstract

**Background:** Breast cancer is a complex disease that affects millions of people and is the leading cause of cancer death worldwide. There is therefore still a need to develop new tools to improve treatment outcomes for breast cancer patients. Electronic Health Records (EHRs) contain a wealth of information about patients, from pathological reports to biological measurements, that could be useful towards this end but remain mostly unexploited. Recent methodological developments in deep learning, however, open the way to developing new methods to leverage this information to improve patient care.

**Methods:** In this study, we propose M-BEHRT, a Multimodal BERT for Electronic Health Record (EHR) data based on BEHRT, itself an architecture based on the popular natural langugage architecture BERT (Bidirectional Encoder Representations from Transformers). M-BEHRT models multimodal patient trajectories as a sequence of medical visits, which comprise a variety of information ranging from clinical features, results from biological lab tests, medical department and procedure, and the content of free-text medical reports. M-BEHRT uses a pretraining task analog to a masked language model to learn a representation of patient trajectories from data that includes data that is unlabeled due to censoring, and is then fine-tuned to the classification task at hand. Finally, we used a gradient-based attribution method -to highlight which parts of the input patient trajectory were most relevant for the prediction.

**Results:** We apply M-BEHRT to a retrospective cohort of about 15 000 breast cancer patients from Institut Curie (Paris, France) treated with adjuvant chemotherapy, using patient trajectories for up to one year after surgery to predict disease-free survival (DFS). M-BEHRT achieves an AUC-ROC of 0.77 [0.70-0.84] on a held-out data set for the prediction of DFS 3 years after surgery, compared to 0.67 [0.58-0.75] for the Nottingham Prognostic Index (NPI) and for a random forest (p-values = 0.031 and 0.050 respectively).

In addition, we identified subsets of patients for which M-BEHRT performs particularly well such as older patients with at least one lymph node affected.

**Conclusion:** In conclusion, we proposed a novel deep learning algorithm to learn from multimodal EHR data. Learning from about 15 000 patient records, our model achieves state-of-the-art performance on two classification tasks. The EHR data used to perform these tasks was more homogeneous compared to other datasets used for pretraining, as it exclusively comprised adjuvant treated breast cancer patients. This highlights both the potential of EHR data for improving our understanding of breast cancer and the ability of transformer-based architectures to learn from EHR data containing much fewer than the millions of records typically used in currently published studies. The representation of patient trajectories used by M-BEHRT captures their sequential aspect, and opens new research avenues for understanding complex diseases and improving patient care.

## 1 INTRODUCTION

Breast cancer is by far the most commonly diagnosed cancer among women (almost 2.3 million cases worldwide in 2022) and the leading cause of cancer death worldwide (1).

Among the various treatment options, adjuvant chemotherapy is proposed to patients after first-line surgery to lower the chance that the cancer will return. It is a widely used treatment option, and is offered in many cases, unless the tumor was small, did not show sign of aggressiveness, and no lymph nodes were affected. However, recurrence or death are still possible. Accurately identifying the patients most likely to relapse is therefore important to inform both treatment selection and future research to propose better therapeutic options.

One of the most commonly used prognostic tools for breast cancer is the Nottingham Prognosis Index (NPI), which uses a combination of three clinical features (tumor size, tumor grade, and number of lymph nodes) and was proposed in 1982 (2). Since then, many authors have used statistical and machine learning algorithms to build breast cancer relapse predictors from clinical features; however NPI still seems to be the most robust criterion (3), despite its limitations.

In the quest for improving the future outcome of patients, there has been a growing interest over the years for including information besides clinical features into prognostic tools. These modalities include biological measurements (4), magnetic resonance imaging (5), ultrasound images (6), histopathological images or gene expression data (7). The papers cited show that combining different modalities improves prediction performance.

However, these modalities are not always available for all patients treated. For this reason, other authors have taken advantage of the considerable information present in medical reports that constitute the EHR of patients, using named entity recognition techniques to extract relevant terms from clinical notes (8, 9).

Among those, transformer-based models inspired by BERT (Bidirectional Encoder Representations from Transformer) (10), an architecture that has significantly outperformed previous methods on a large variety of natural language processing tasks and continues to drive advancements in the field, have recently gathered a lot of interest. Their superiority is explained by the use of self-supervised pretraining tasks, such as masked language modeling and next sentence prediction, which allows them to learn better representations of the data. These architectures have been successfully transposed to patient trajectories by seeing them as sequences of medical events rather than of words (11, 12, 13, 14, 15). To the best of our knowledge, however, none of these have considered cancer-related clinical outcomes, possibly because they are typically applied to very large cohorts of millions of patients.

In this paper, we present several new transformer architectures for predicting clinical outcomes from multimodal EHR data, which consider patient trajectories as sequences of medical visits represented by both tabular data (clinical features, biological measurements, therapies, nature of the visit) and free-text medical reports. We evaluate our proposed method on the prediction of disease-free survival in breast cancer, on a cohort of several thousands of patients. We pretrain the models on the equivalent of a masked language model, which can also be trained on records excluded from the classification training set because they were censored.

## 2 MATERIALS AND METHODS

### 2.1 Data

In this work, we used data extracted from the EHR system from Institut Curie in Paris (France). All data collected were pseudonymized. Additionally, individuals under 18 years of age, with a history of previous cancer, under guardianship, or unable to provide consent were excluded from this study. Every patient included in the study has completed and signed a research informed consent form. The study was approved by the Breast Cancer Study Group of Institut Curie and was conducted according to institutional and ethical rules concerning research on tissue specimens and patients.

We built a data base of 15 150 unique patients, treated with adjuvant chemotherapy for breast cancer between 2005 and 2012. The data base contains general descriptors of patients (such as age, sex, or weight) as well as information about each visit in their medical record: clinical information such as tumor size or cancer subtype, biological markers (tumor markers, counts of leukocytes and their subtypes) if they were measured, treatment information, and free-text notes. Finally, the patients are annotated with survival and recurrence information.

Free-text notes are unstructured narrative descriptions or notes entered by healthcare professionals. Unlike the structured data, which is organized into predefined fields, free text allows healthcare providers to input progress reports and relevant patient information recorded during patient journey, in a more natural manner. Free text reports from cytopathology or radiology also capture key information from medical images, as captured by experts. Those medical reports comprise free-text clinical notes for consultations, as well as free-text reports of cytopathology, radiology, surgery, and blood tests. All reports are written in French.

### 2.2 Preprocessing

#### 2.2.1 Tabular data preprocessing

We first describe how we processed the structured or tabular, a.k.a structured, data describing each medical event for each patient.

##### 2.2.1.1 Biological measurements

From biological measurements, we only kept features that have less than 30% of missing values: MONO, LEUK, LYMP, PN and CA 15-3. All numerical values have to be discretized to enable tokenization. We binarized biological measurements into two values: 1 if the value is outside the normal range for the biological measurement, and 2 otherwise. Figure S1 in the Supplementary Material shows the distribution of biological measurements; the medical normal range of these biological features can be found in Table S1 in the Supplementary Material.

In addition, we also computed the differences Δ*_t_* = *v_t_ − v_t−_*_1_ between the current visit’s biological value *v_t_* and the previous visit’s value *v_t−_*_1_. We then discretized the Δ values by dividing them by ten and rounding. This captures more subtle variations in biological measurements evolution than the mere abnormal/normal values.

##### 2.2.1.2 Clinical information

From the clinical information, we included both longitudinal and non-longitudinal features: age, undergone therapies, and tumor size on the one hand, tumor grade and number of nodes involved at diagnostic as well as breast cancer molecular subtypes (Luminal, TNBC, HER2+/RH-, HER2+/RH+) on the other. Age is computed at each visit and discretized by rounding to the nearest integer. Descriptive statistics of the age, breast cancer subtype, grades, number of lymph nodes involved, tumor size and biological measurements are given in Table S1 in the Supplementary Material.

We combined tumor size, tumor grade and the number of lymph nodes involved into the NPI (2), a commonly used, clinically relevant and robust prognostic tool (3). The NPI is computed as NPI = 0.2 *×* tumor size (cm) + tumor grade + lymph nodes stage, where the lymph nodes stage is computed as 1 (0 nodes), 2 (1 to 3 nodes) or 3 (*>* 3 nodes). The lower the score, the higher the chance of survival 5 years after surgery. The tumor size is measured at various points in the cancer journey. We kept for this study the clinical tumor size assessed at diagnosis when the tumor is palpable, and the pathological tumor size which is the histological size of the tumor extracted at the surgery. The NPI is recalculated with each new tumor size measurement, hence termed as the dynamic NPI (dNPI). For patients with at least one available feature among the three required for calculating the dNPI, we imputed missing tumor sizes using the mode value among samples of the same clinical or pathological tumor stage (TNM) status. The number of involved lymph nodes is the sum of the number of affected sentinel nodes and axillary nodes. We imputed missing number of nodes to zero and missing tumor grade to G2 (grade 2), based on the most frequent values in our data. The higher the dNPI, the lower the chance of survival.

Following Blamey et al. (16), we categorized dNPI into six prognostic groups (PG): Excellent (EPG) (NPI *≤* 2.4), Good (GPG) (2.4 *<* NPI *≤* 3.4); Moderate I (MPG I) (3.4 *<* NPI *≤* 4.4), Moderate II (MPG II) (4.4 *<* NPI *≤* 5.4), Poor (PPG) (5.4 *<* NPI *≤* 6.4) and Very Poor (VPPG) (NPI *>* 6.4).

Because M-BEHRT can handle missing values (see Section 2.3.1), we did not impute missing values for longitudinal features. However, for the baselines, we opted to impute the tumor size, number of nodes, grades and cancer subtype by an aberrant value of 999. Using an aberrant value allows the model to explicitly identify and differentiate imputed values from the actual data, by analogy with not locating a token within a sentence when using M-BEHRT.

##### 2.2.1.3 Therapies, department and procedure

Therapies are inferred by considering the occurrence date for the surgery, the start and end dates for hormone-therapy, chemotherapy and anti-HER2 treatment, and the number of doses administered for the radiotherapy. This inference incorporates the therapeutic protocol of Institut Curie (see Figure S2 in the Supplementary Material). Subtherapies, also inferred from this protocol, provide additional information about the specific molecules given in the case of chemotherapy or anti-HER2 therapy, radiation types in the case of radiotherapy, and specific surgical procedures including both breast and axillary surgeries. A list of all possible values for the therapies and subtherapies fields is given in Table S3 in the Supplementary Material.

Finally, medical visit department and procedure names are available within the headers of free-text reports. We normalized department and procedure names by removing accents, punctuation and special characters. We merged synonyms into a single word: for example, *anapath*, *anatomopathologie* and *anatomo-cyto-pathologie* are merged into *anatomo-cyto-pathologie* (anatomical cytology in English). To do so, we sifted through the corpus vocabulary, identifying and unifying synonyms and/or differently written terms to enhance coherence of the medical history. We also removed words that appear fewer than 100 times in the whole corpus.

##### 2.2.1.4 Disease-Free Survival at 3 years

Finally, we defined a binary classification task by labeling each patient with whether they had survived disease-free 3 years after the surgery.

We retained patient history up to one year after first surgery and starting from 6 months before the breast cancer diagnosis. This choice of one year after the first surgery as an index date ensures that we use as much of the patient’s history as possible, without capturing an actual relapse. We removed patients who relapsed before the index date, as well as patients censored before 3 years after the first surgery, as depicted in Figure 1. All patients had at least 3 visits in their medical history. This results in 8 089 patients, with 6.2% having a negative disease-free survival (DFS) status.

**Figure 1.**
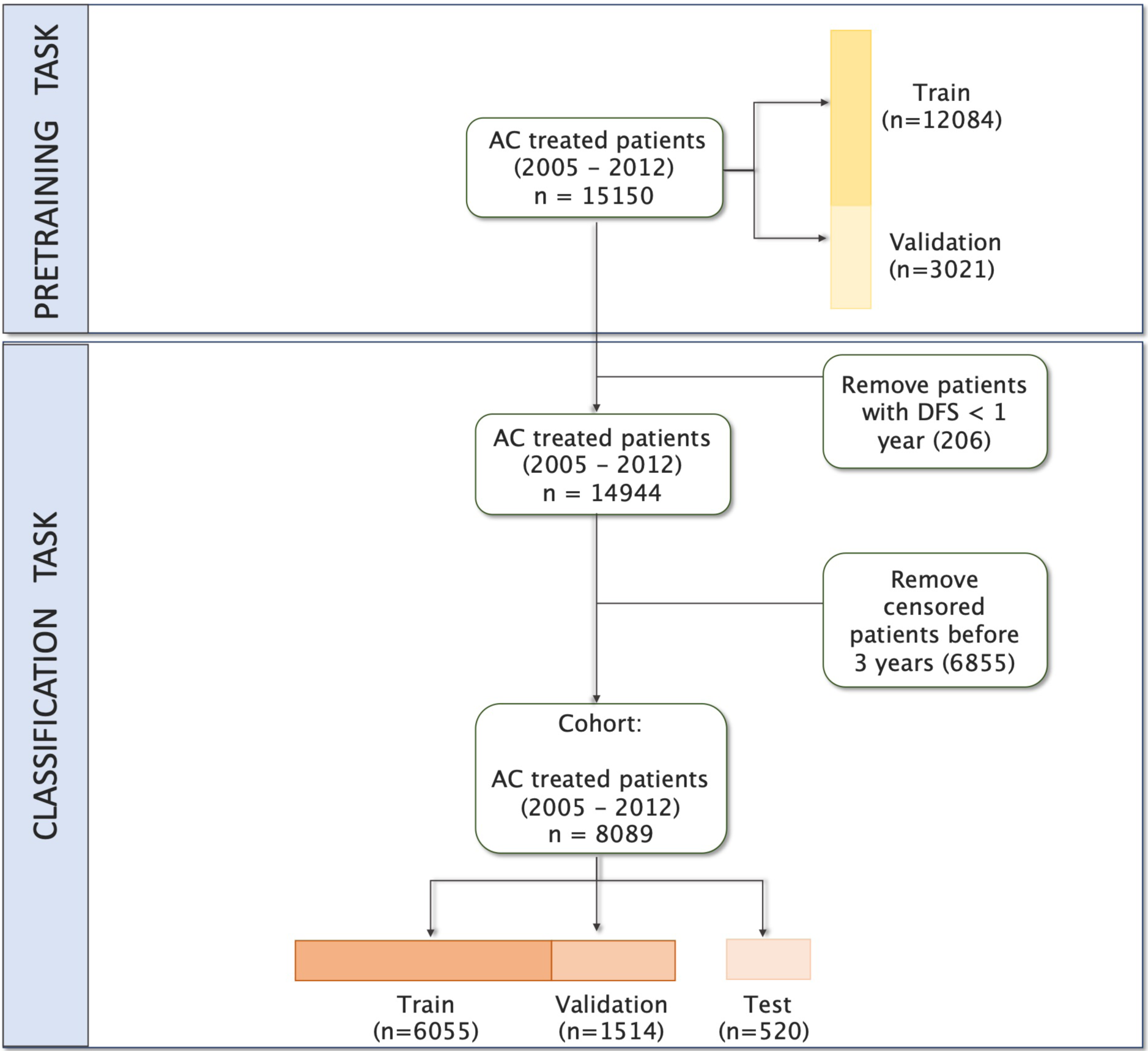
Flowchart of study inclusion and exclusion.

For the evaluation of our models, we held out a test set containing 520 patients, with a proportion of negative samples (6.1%) similar to that of the whole data set. For pre-training tasks requiring no labels (see Section 2.3.2), we used all patients and their full history.

#### 2.2.2 Free-text reports preprocessing

Free-text reports represent unstructured textual descriptions of medical information recorded by medical experts. They can be clinical notes, that is to say, information recorded during patient encounters with clinicians, or reports made by specialists (laboratory biologists, radiologists, histopathologists) to interpret the results of medical exams. The average number of visits, reports, and words per report in our data are given in Table S1 in the Supplementary Material.

Unlike tabular data, that is recorded in a standardized way at least within a hospital, medical reports are highly variable, as they allow each healthcare provider to be distinctive in format, style, or terminology. Moreover, the semantic related to the medical field is complex, using abbreviations, acronyms, and medical jargon (17). Therefore, in addition to common NLP preprocessing steps (normalization, removal of noisy entities, adverbs, stopwords and text delimiters), our text preprocessing pipeline includes steps that are specific to medical reports. The full text preprocessing pipeline is described on Figure S3 in the Supplementary Material, and we describe in Text S1 in the Supplementary Material the steps that are specific to clinical text.

### 2.3 Multimodal BEHRT

Information retrieved from EHR are generally time stamped events. In this study, this information is organized as structured or tabular data (for numerical values) collected over time, along with a series of free-text medical reports throughout the patient’s journey. As in Natural Language Processing, EHR can be transformed into sequences of tokens, where each token represents a unit of information from the EHR rather than a linguistic unit. These sequences can then be fed into language models such as transformers (18). This was first proposed by Li et al. (11), who introduced BEHRT (BERT for EHR), an architecture based on that of BERT (Bidirectional Encoder Representations from Transformers) (10) to predict future conditions from a sequence of diagnoses.

Here we propose Multimodal-BEHRT (M-BEHRT), which combines two transformer-based deep learning models of architecture inspired by BEHRT’s: Tabular BEHRT and Text BEHRT. Tabular BEHRT considers that each medical visit is described using structured data: the department in which it took place, the corresponding procedure, as well as clinical and biological measurements available at this time. Like BERT and BEHRT, Tabular BEHRT combines a pre-training task (Masked Language Model) with a downstream task (the classification task), but applies it to a multimodal EHR tabular dataset. Text-BEHRT considers that each medical visit is represented by a free-text medical report. Text BERT uses adapted pretrained embeddings to build a sequence that serve as input for the classification task. M-BEHRT is a meta-model that combines Tabular BEHRT and Text BEHRT through a cross-attention module (19).

In what follows, we first describe how we construct patient trajectories (Section 2.3.1 from multimodal tabular data (Section 2.3.1.1) as well as free-text reports (Section 2.3.1.2). We then describe in Section 2.3.2 the self-supervised approach used for learning embeddings of multimodal patient trajectories, and in Section 2.3.3 the architecture we propose for the binary classification of patient trajectories. Finally, we describe the baselines used to evaluate our models in Section 2.4.

#### 2.3.1 Multimodal sequence construction

##### 2.3.1.1 Patient trajectory representation from structured data

By analogy with Natural Language Processing data, a patient’s history can be seen as a document, where visits serve as sentences, and the events within the visits act as tokens. In our final data, the medical sequence consists of a sequence of visits that are chronogically ordered.

We used dates from the medical reports to construct medical chronological sequences. Each visit is described by the specific department and procedure from which the report originates, which contextualizes additional features, which are incorporated as available.

As illustrated on Panel A of Figure 2, each visit is therefore described by at most 17 features: biological measurements that include binary values and deltas of measurements of the 5 biological markers, the medical department where the visit took place, the type of procedure the visit corresponded to, the therapy and sub-therapy administered, the patient’s age, the dNPI and the breast cancer subtype (which is static but repeated at each visit).

**Figure 2.**
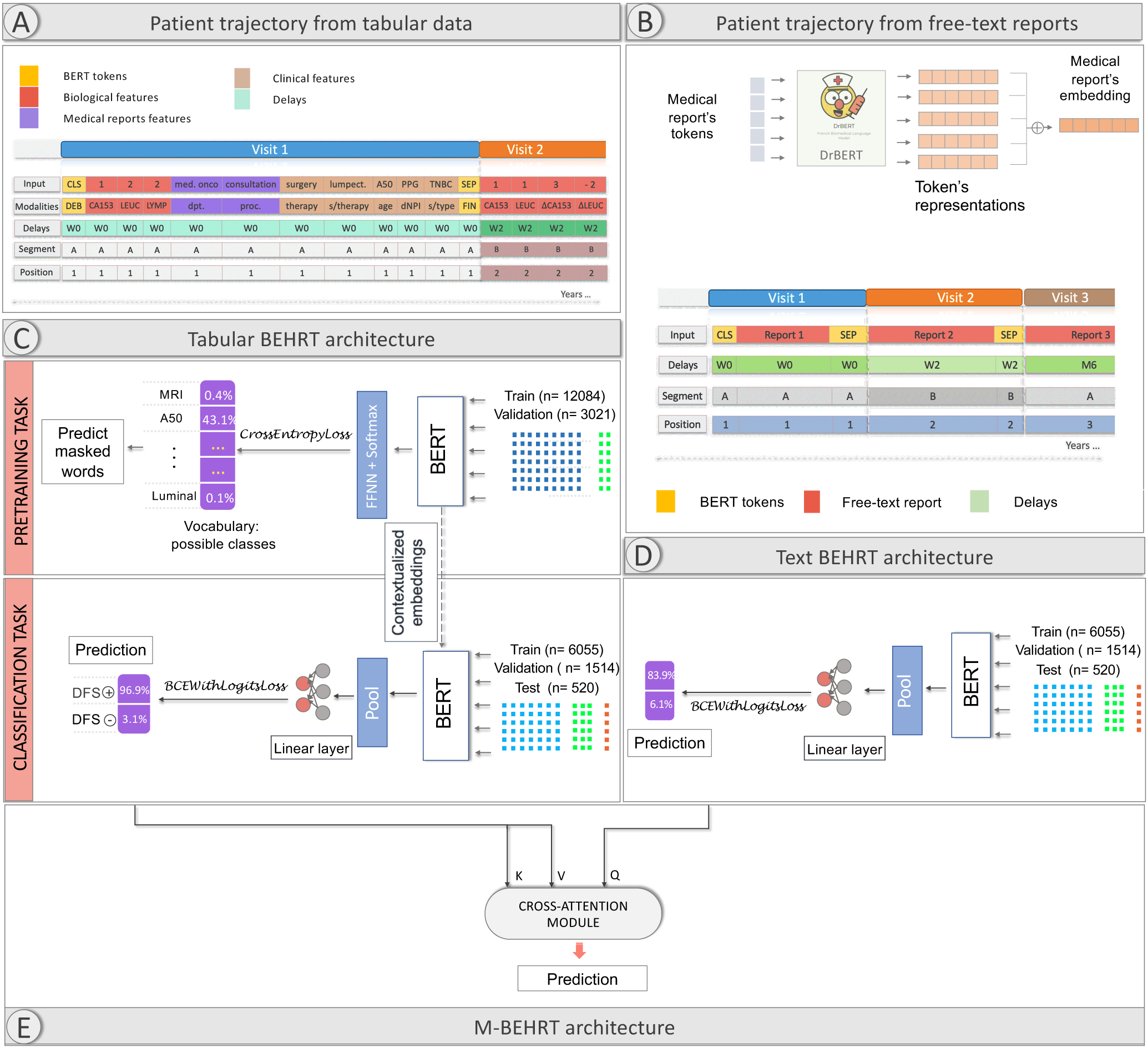
M-BEHRT architecture. Panel A: representation of patient trajectory using tabular data. Panel B: representation of patient trajectory using free-text reports. Panel C: architecture of Tabular BEHRT (learning from patient trajectories represented from tabular data as in Panel A). Panel D: architecture of Text BEHRT (learning from patient trajectories represented from free-text reports as in Panel B). Panel architecture of M-BEHRT, learning from both representations by combining Tabular BEHRT and Text BEHRT with cross attention.

A separate modality layer indicates what kind of feature each measurement corresponds to. Generally speaking, this could be set to simply indicating the modality (biological, clinical, visit), but here we chose to be specific and encode the feature name. This allows us in particular to deal with missing values, which can simply be skipped as the modality layers provides the information of what feature is at each position. The modality layer allows the algorithm to treat each modality differently.

As in BERT and BEHRT, a sequence of visits starts with the special token CLS, and visits are separated with the special token SEP.

Whereas BEHRT captures temporal information by including the age of the patient in a separate layer, we kept age as other clinical descriptors in the main input layer, but added another special embedding layer that represents the delay between the next visit and the previous. We discretized delays, as in Pang et al. (12), into W0-3 (under 1, 2, 3, or 4 weeks) for delays shorter than 4 weeks, M1-12 (under 1 month up to under 12 months) for delays shorter than a year and LT (long term) for delays longer than a year.

One of the notable constraints in BERT-like models is token capacity: they process tokens in fixed-size sequences of at most 512 tokens. While this size is arbitrary and varies depending on the exact BERT architecture and implementation, it cannot take much larger values, as it is linked to the memory usage of the self-attention mechanism of BERT, which grows quadratically with the number of tokens (each token being attentive to every other token). There is therefore a tradeoff between the number of features/tokens used to describe each visit, and the number of visits that can be considered. This is alleviated by the exclusion of both missing values and biological delta values equal to zero (corresponding to an absence of change in measurement), which is possible as the modality layer informs the architecture as to the kind of feature each token corresponds to. In practice, if the patient trajectory still exceeds 512 tokens, we only consider the first 512 tokens, which represent the initial interactions of the patient with the healthcare system, and inform about initial diagnostic visits and treatment decisions. Figure S4 in the Supplementary Materials shows how much information is excluded from patient trajectories due to restricting data to the 512 first tokens.

Panel A of Figure 2 illustrates this representation of patient trajectories based on tabular data.

##### 2.3.1.2 Patient trajectory representation from free text

In addition, we assume that important information is contained within the text itself of the free-text reports. We therefore build a sequence of free-text reports, ordered chronologically from the date of the diagnosis until the index date (one year after the first surgery). As shown in Table S2 in the Supplementary Material, the number of reports per patient and the length of each report are such that these create very long documents (on average 34 reports, averaging 159 words each, for a total of more than 5 000 words per patient history). However, while BERT has proven to be highly effective in capturing contextual relationships and semantic nuances in text, it can only process sequences of at most 512 tokens, due to the memory footprint of the self-attention mechanism.

This constraint again poses challenges when dealing with lengthy documents such as a sequence of medical reports (20). Using transformers to classify long documents is still a topic of open research (21). The most straightforward approach consists in truncating inputs to fit within the allowed number of tokens, typically by using the first, last or middle tokens. However, limiting patient history to 512 tokens may result in major information loss and hence produce incomplete representation of medical reports. Other approaches such as Big Bird (22) or Nyströmformer (23) use sparse or low-rank approximations of the self-attention matrices. However, existing pretrained models typically do not handle more than 4 096 tokens, which is still too short for some of the patients in our data set. In addition, they have only been trained on English corpora whereas our medical notes are in French. Nevertheless, our corpus is much too small to train a transformer model from scratch. Finally, many approaches consist in dividing long text into chunks smaller than 512 tokens and combining their embeddings, whether through an additional layer of self-attention in a hierarchical model (24) or by pooling (25). In the absence of a clear consensus on which of these strategies is likely to perform best (21, 25), we chose here to use a simple aggregation strategy. More specifically, we construct the embedding of every report by summing the embeddings of all tokens it contains, and construct sequences not of token embeddings, but of reports embeddings.

We obtain token embeddings from DrBERT (26), a state-of-the-art transformer model, based on the RoBERTa architecture (27) and trained on a French biomedical corpus which contains 7GB of clinical data from multiple sources. We can then train a BERT model on the sequences of reports embeddings. To account for temporality, we add an embedding layer of delays between reports. Finally, we use BERT special tokens: CLS for the start of a medical history and SEP to separate reports from different visits. This representation is illustrated on Panel B of Figure 2.

#### 2.3.2 Pretraining task

To improve the embeddings of patient trajectories built from structured data, we follow the example of BEHRT and pre-train a Masked Language Model (MLM) on the representations described in Section 2.3.1.1.

As in Natural Language Processing, the MLM is designed to predict missing or masked tokens within a patient’s history, using the bidirectionaly context provided by the surrounding tokens. Its goal is to learn contextual representation of the medical events in the patient’s history. For this purpose, in this pre-training phase Tabular-BEHRT uses the whole cohort of 15 150 patients and the entire sequence of events for each patient, from the date of diagnosis to the date of death or censorship, with a length average of 506(*±*466) tokens. We randomly replaced 15% of the tokens with a special MASK token. We swapped another 2% with another token at random; this adds a limited amount of noise, encouraging the model to learn a more robust and generalizable representation of patient trajectories. As shown on Panel C of Figure 2, the MLM part of M-BEHRT is a transformer-based architecture that generates probabilities for each token in the vocabulary, computed using softmax over the model’s output logits, as a multilabel learning task.

We first split the dataset into a training (90%) and a validation set (10%) in order to prevent overfitting. Then, all the embeddings from the training set are randomly initialized and fed to the MLM. We use Bayesian optimization to find the best set of hyperparameters, with precision as a criterion. For robustness, we run the model five times with five different random seeds for the sequence masking, and use as final token embeddings for the downstream classification tasks the mean values of standardized embeddings from these five runs.

The pretraining task solely concerns tabular data, to establish effective representations of tabular events within the patient trajectory. For text data, running an MLM on the whole medical corpus would require more computational resources than available.

#### 2.3.3 Binary Classification

We now describe the architecture of M-BEHRT, a deep neural network to learn binary classifiers from patient trajectories. M-BEHRT is the combination of two architectures: Tabular BEHRT, which learns from patient trajectories built from structured data; Text BEHRT, which learns from patient trajectories built from free text.

Tabular BEHRT consists in using labeled data to fine-tune for classification the network obtained by pre-training on patient trajectories built from structured data. As shown on Panel C of Figure 2, only the last layer is different between pre-training and fine-tuning: here the patient history embeddings are fed to a single feed-forward layer with sigmoid activation.

The architecture of Text BEHRT is illustrated on Panel D of Figure 2. It is again a transformer-based model, which uses report embeddings obtained through the aggregation of DrBERT embeddings as described in Section 2.3.1.2. The same sampling strategy as the one depicted in the previous section is used for this task.

Finally M-BEHRT combines information from tabular data and free-text reports by integrating Tabular BEHRT and Text BEHRT using a cross-attention module(19). The cross-attention module extends the capabilities of traditional transformer architectures to handle multiple data modalities in a unified framework. Hence M-BEHRT is expected to harness the complementarity of the information encoded in different modalities to improve predictive power.

As shown on Panel E of Figure 2, logits from structured data trajectories and the text trajectories are computed using their respective models. The cross-attentions layer calculates attentions with the logits as key, value and query. Logits from Text BERHT used as query interact with logits from Tabular BERHT that represent key and value. The loss is backpropagated to the cross-attention module. To do so, logits must have same size. Therefore, logits from Text BEHRT are first fed through a single feed-forward layer to obtain an embedding of the same size as logits from Tabular BEHRT.

In contrast, cross-attention is used when there are two distinct sets of inputs. One set of inputs (the “query”) interacts with another set (the “key” and “value”). The model attends to the “key” sequence to inform the processing of the “query” sequence. This is commonly used in models where input data needs to interact, such as translating a sentence in one language to another.

Because the labeled data is typically imbalanced, we implemented a stratified batches strategy, which consists in loading the same proportion of positive and negative samples for each batch, with replacement for the positive instances (the minority class). This sampling strategy allows us to train on balanced batches.

### 2.4 Comparison baselines

To evaluate our models, we developed several comparison baselines. The first is the NPI measured at the date of diagnosis, a tool that is currently used in the clinic to predict prognosis. In addition, we developed baselines using classical machine learning methods: random forests classifiers (RF), logistic regression (LR), and support vector machines (SVM). These machine learning models (RF, LR and SVM) use the same input data as M-BEHRT, but cannot directly use sequential information. For dynamic tabular data (procedure name, department name, binarized biological measurements), sequences of events are transformed into number of occurences of events. Clinical features (age, therapies, tumor size, tumor grade, breast cancer molecular subtype and number of nodes) are kept static, using their values at the time of diagnosis. Regarding free-text reports, we created a table where each feature of the report embeddings (of 768 dimensions) becomes a column. We imputed missing values with zero (0) for both of the inputs. For M-BEHRT, outputs from tabular data baselines and from text data baselines (specifically their logits) constitute inputs to a secondary model (meta-model) which makes the final prediction.

In order to consider class imbalance and prevent the model from being biased towards the majority class, we choosed the strategy of assigning different weights to each class during training. These weights are inversely proportional to class frequencies in the training data. By penalizing the majority class, the model is ensured to have enhanced performance on minority classes.

### 2.5 Model selection

For model selection, we split the training data (8 289 patients, excluding the held-out data set of 520 patients) into a training and a validation sets (respectively 90% and 10% of the data). For each method, we use Bayesian optimization (28) to find the optimal set of hyperparameters, using the Average Precision Score (APS) on the validation set as a performance criterion.

### 2.6 Computational resources

We used Python to code models and analyses pipelines for this study, in particular scikit-learn (29) for the classical machine learning models, hyperopt (28) for Bayesian optimization, spaCy (30) for natural language processing tasks, and PyTorch (31) for the implementations of Tabular BEHRT, Text BEHRT and M-BEHRT, which are built on that of BEHRT (11). The masked language model and DFS classification model were computed on NVIDIA A40-46GB Graphical Processing Units (GPU).

## 3 RESULTS

### 3.1 Patient trajectory embeddings

#### 3.1.1 Tabular patient trajectory embeddings

We first focus on the Masked Language Model (see Section 2.3.2) and evaluate the quality of the patient trajectory embeddings learned during the pre-training phase of Tabular BEHRT.

The optimal hyperparameters we identified for the MLM are 5 hidden layers with 12 attention heads, a hidden size of 144, an intermediate layer size of 133, a training duration of 120 epochs, using Adam optimizer with a learning rate set to 1e-3 and a batch size of 64.

To assess the performance of the MLM, we ran the model five times with five different random seeds for the sequence masking. We also compute a baseline by running the MLM on a data set in which tokens have been randomly reordered within each sequence. This approach disrupts the inherent sequential structure of the data, and creates a scenario where the model should not be able to rely on contextual relationships between tokens. Hence, comparing the MLM’s performance on shuffled sequences against its performance on original sequences offers a benchmark for assessing the impact of contextual information on the model’s predictive capabilities. The precision of these models (proportion of correctly predicted masked tokens) on the held-out validation set is shown on Figure S5 in the Supplementary Material.

The MLM is able to predict masked tokens with a precision of 72% on the validation set, a performance that is not significantly different from the one on the training set, highlighting the absence of overfitting. In addition, this precision is significantly higher than the precision of 55% obtained when shuffling the sequences, which shows that the MLM does indeed capture contextual information. We also note that the precision of the MLM of BEHRT reported by Li et al. (11) on sequences of diagnoses is of 66%. While it is difficult to compare this performance to ours due to the different nature of the tasks, it indicates that the MLM provides embeddings of sufficient quality to perform supervised learning in a second stage.

We further evaluate embeddings generated by the MLM by visualizing token embeddings through two-dimensional plotting along the first two components of a t-distributed Stochastic Neighbor Embedding (t-SNE) as shown on Figure 3. This figure shows how the MLM capture semantic relationships between tokens and contextual information. Tokens belong to the same modality (therapies, variation in biological features, breast cancer subtypes) tend to cluster together, with the exception of procedures and departments, which tend to be mixed together. This is however unsurprising, as some procedures and departments are tightly linked; for example, panel F shows that the embedding of the “nuclear medicine” service is quite close to the embeddings of “radiology”, “scanner” and “MRI” procedures, while panel D shows that the embedding of the “radiotherapy” service is quite close to the embeddings of several procedures all relating to the proposal, prescription, initiation, unfolding and ending of treatment by radiotherapy.

**Figure 3.**
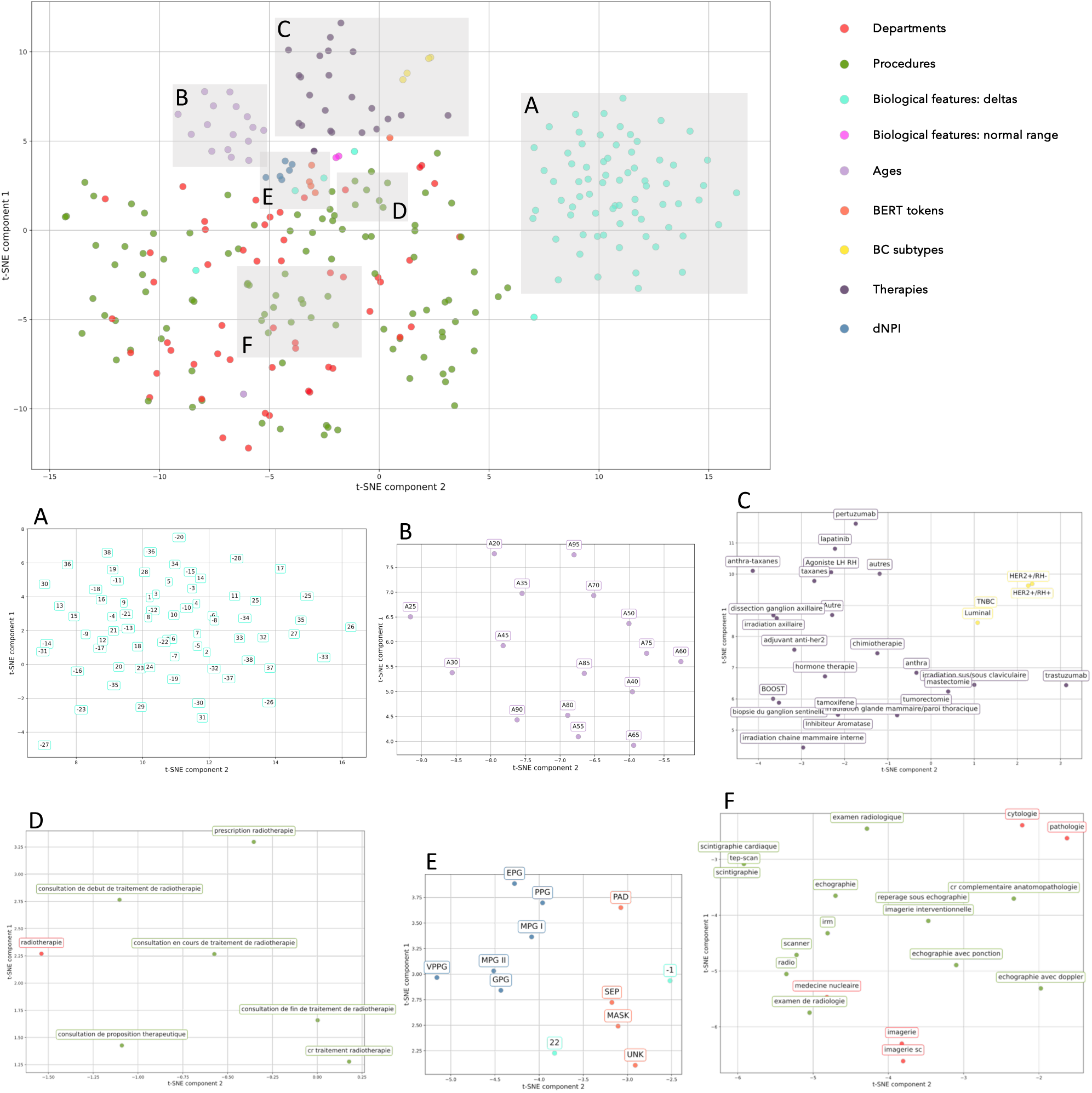
t-SNE of Tabular BEHRT tokens embeddings as learned by the Masked Language Model. Panels A through F zoom in on specific section of the plot. Panel A corresponds to a cluster of deltas in biological measurements. Panel B shows that age tokens cluster together. Panel C shows that therapy token, on the one hand, and breast cancer subtypes, on the other, cluster together. Panel D and F show two different clusters of procedures and departments. Panel E show that dNPI tokens cluster together, as well as BERT special tokens.

#### 3.1.2 Medical reports embeddings

We first evaluate the quality of the medical reports embeddings obtained by pooling tokens embeddings extracted from DrBERT by visualizing them after their projection into a 2D space using t-SNE. The proximity of reports within this space corresponds to their semantic similarity. As shown in Figure 4, this visualization provides a comprehensive overview of the clustering patterns, demonstrating the potential of DrBERT embeddings in representing French medical text data.

**Figure 4.**
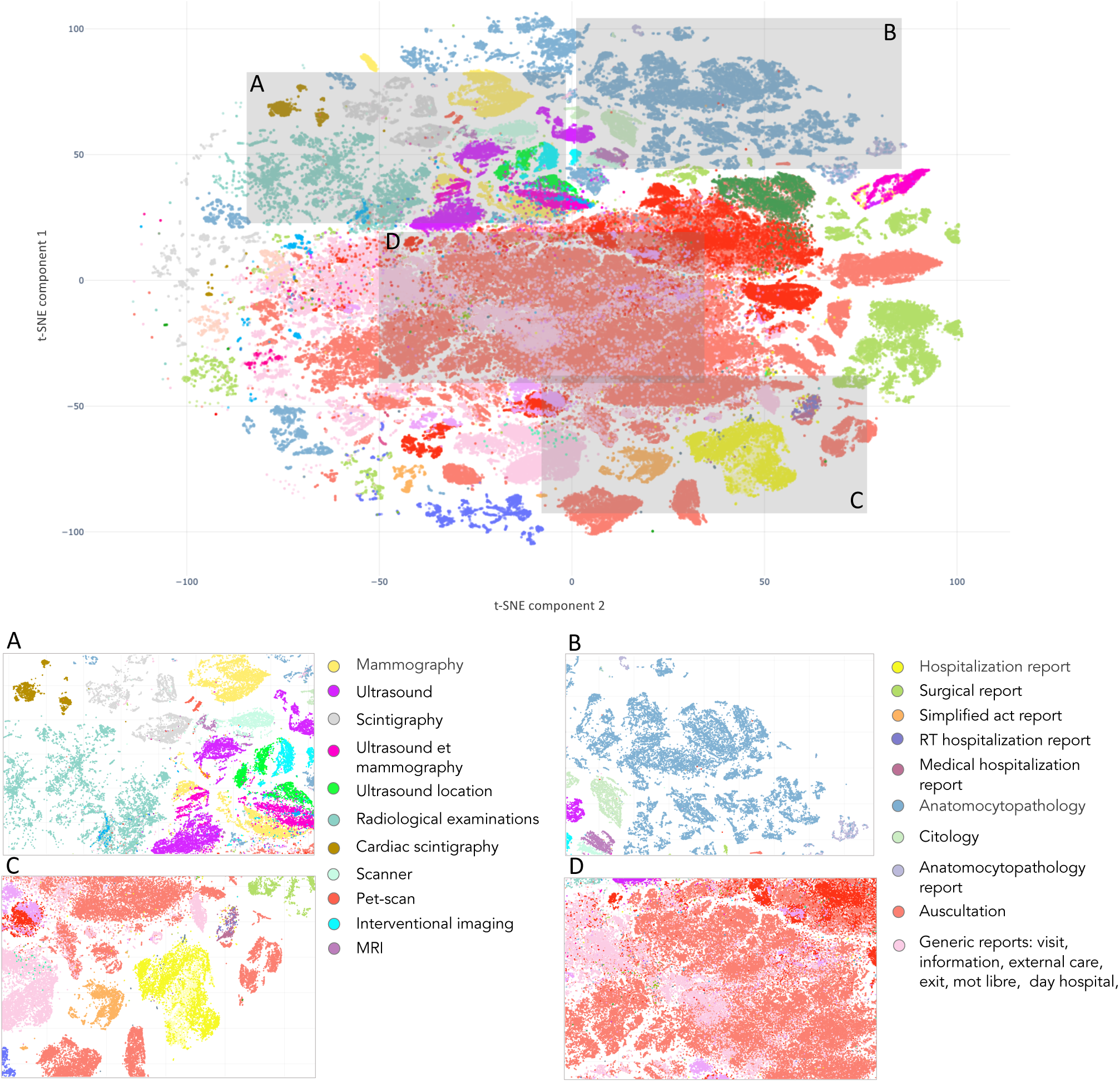
t-SNE of Text BEHRT medical reports embeddings. Each panel correspond to a different departments’ reports with similar information, cluster together.

This figure shows clusters of reports written in the same departments. Additionally, it display promixity between clusters that arise from similar departments. The Panel A groups all reports associated with radiology, including “mammography”, “MRI”, “ultrasound”, or “scintigraphy”. The same pattern is observed in Panel D, which contains the “generic” reports as those related to “discharge”, “external care” or “information”, and in Panel B, with clusters relating to cytology (“anatomocytopathology”, “cytology”). Lastly, Panel C displays reports from various departments positioned closely together.

### 3.2 DFS prediction

#### 3.2.1 Comparison of M-BEHRT with baselines

We report on Figure 5 the ROC curves on the test set of M-BEHRT trained with optimal hyperparameters (see Section 2.5; learning rate of 10*^−^*^3^, batch size of 64, Adam optimizer, 6 epochs of training), as well as of the comparison baselines described in Section 2.4.

**Figure 5.**
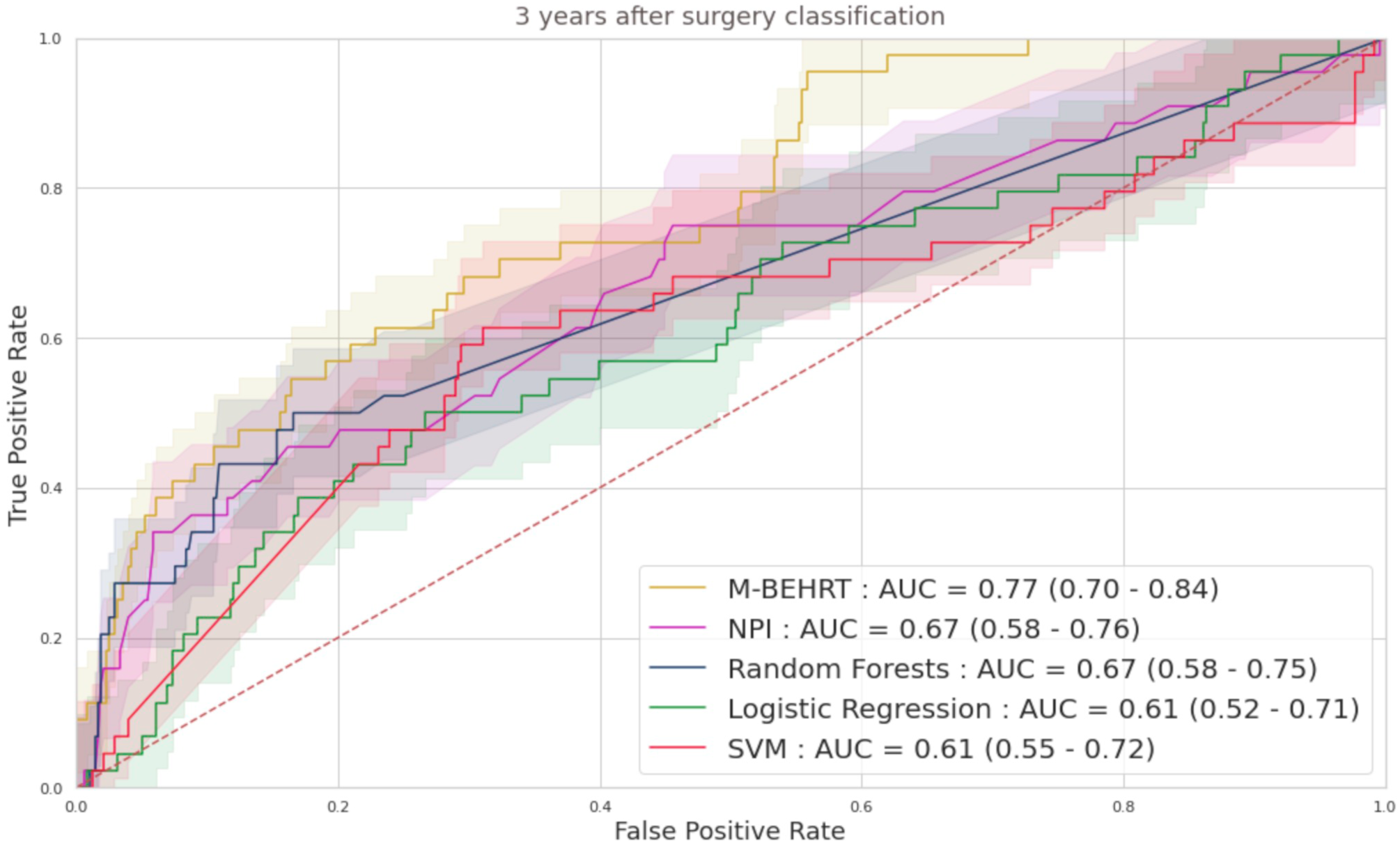
ROC curves M-BEHRT with the baselines for the prediction of disease-free survival 3 years after surgery, on the test set.

Figure 5 shows that all methods perform significantly better than a random classifier (AUC-ROC of 0.5). Moreover, M-BEHRT outperforms all comparison machine learning models.

#### 3.2.2 Ablation study

To better understand the contribution of each modality to the performance of M-BEHRT, we first compared it to the individual performance of its components Tabular BEHRT and Text BEHRT. Figure 6 reports ROC curves for all three approaches, on the test set. The optimal hyperparameters for Tabular BEHRT were a learning rate of 10*^−^*^4^, a batch size of 16, Adam optimizer, and 5 epochs of training; for Text BEHRT they were a learning rate of 5.10*^−^*^4^, a batch size of 32, Adam optimizer, and 99 epochs of training.

**Figure 6.**
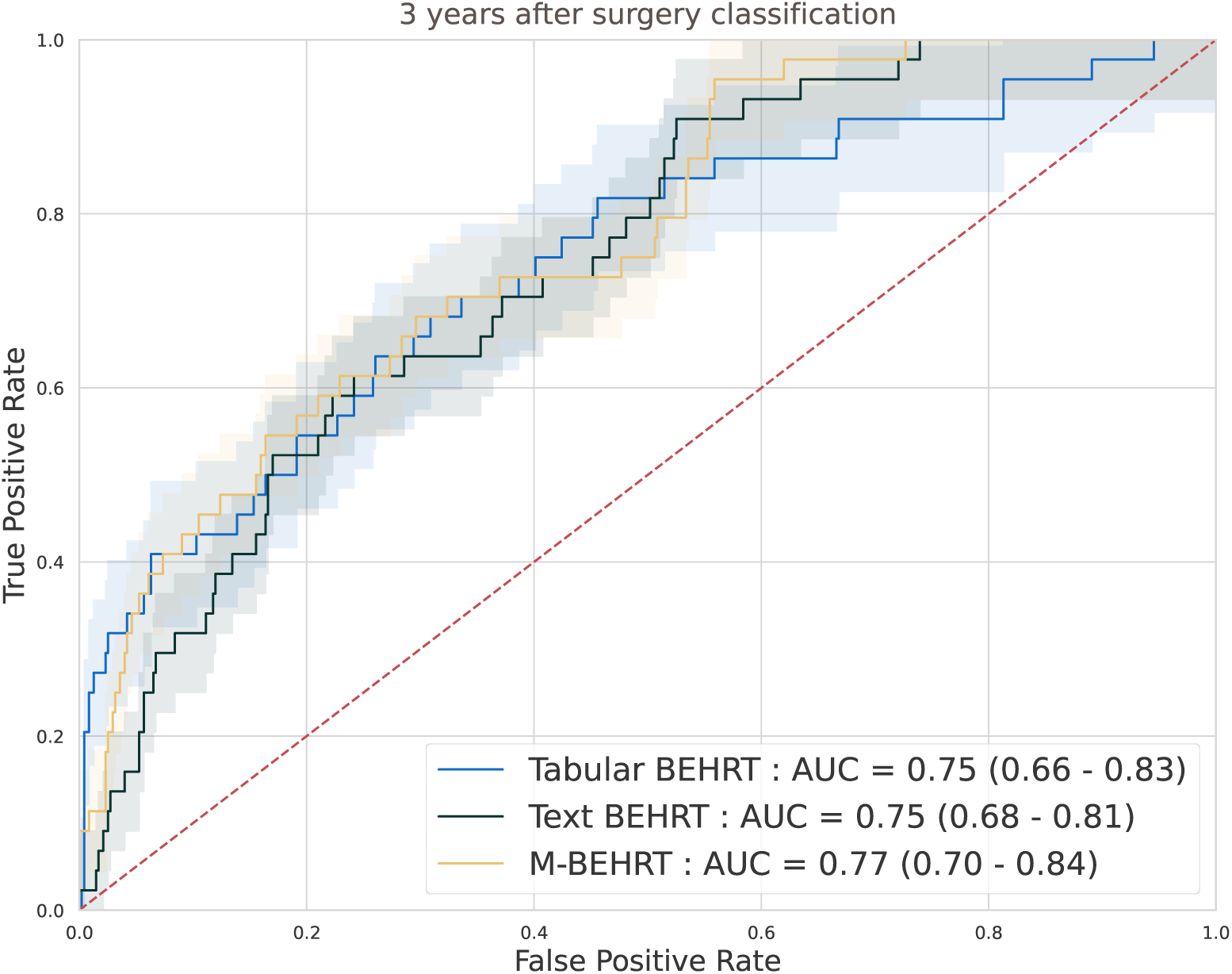
ROC curves comparing Tabular BEHRT and Text BEHRT against their combined model M-BEHRT for the prediction of disease-free survival 3 years after surgery, on the test set.

Although they use different information, Tabular BEHRT and Text BEHRT achieve similar performance on both tasks, highlighting that Text BEHRT can capture relevant information in unstructured medical reports. The combination of both models through cross-attention slightly improves their respective performance, demonstrating the synergistic effect of integrating the strengths of both Tabular and Text BEHRT into a single unified model.

We also performed an ablation study to better understand the contribution of each tabular modality to the performance of Tabular BEHRT. Figure 7 shows the areas under the ROC curves obtained on the test set when removing some of the modalities from Tabular BEHRT. This figures shows that dNPI contributes the most to the performance. However, the addition of the other features, in particular the remaining clinical features (including age and more notably therapies), increases performance substantially. Biological features contribute the least to performance, although they still contain information, as they allow for better-than-random prediction. However, it seems that this information is redundant with that captured by the other features. Performance also drops substantially if information about the nature of the medical visit (department and procedure) is omitted. These observations are consistant across both tasks.

**Figure 7.**
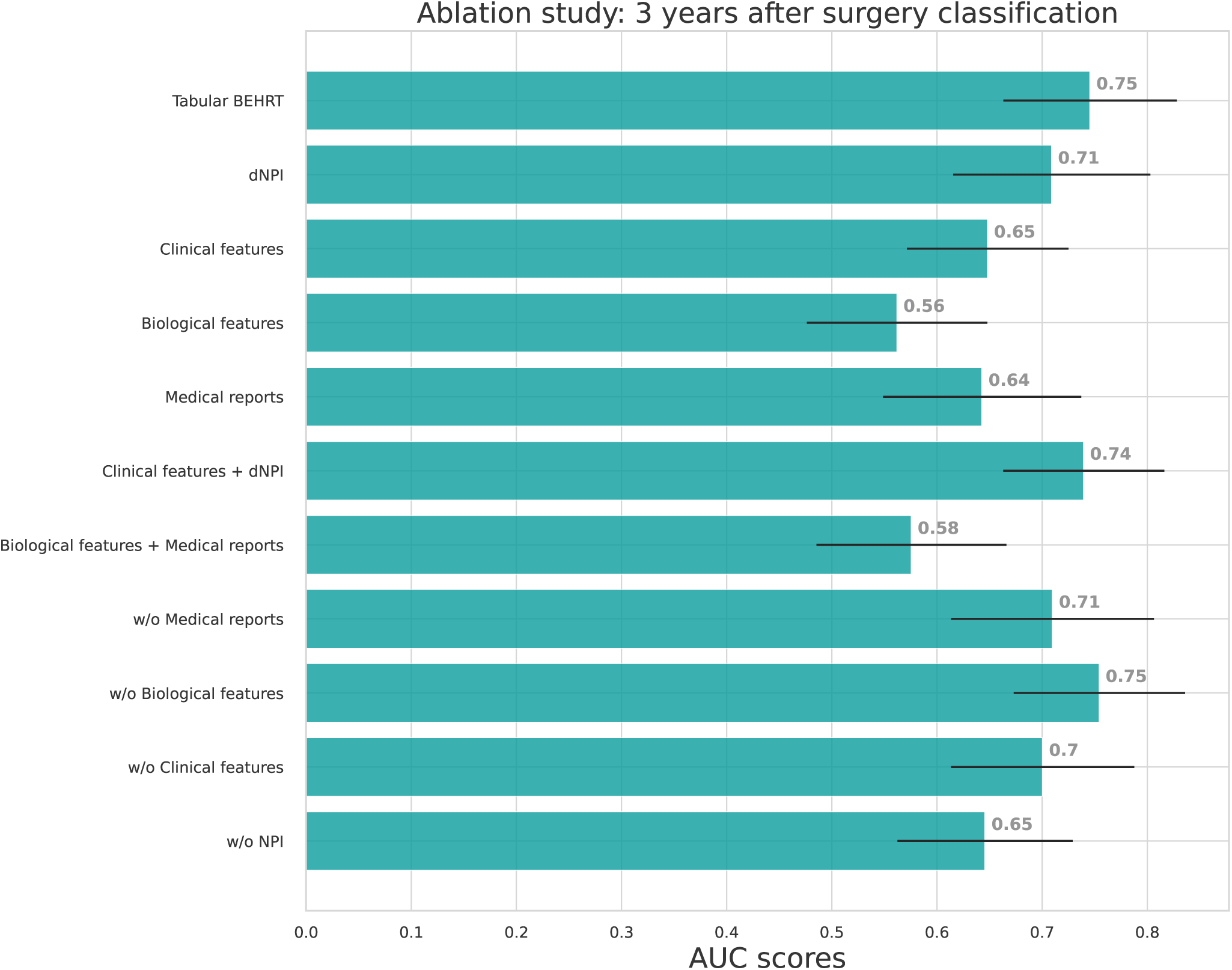
Ablation studies AUC-ROC on the test set for Tabular BEHRT. We present results for the full model (Tabular BEHRT), then using only one of the 4 modalities (dNPI, clinical features, biological features, medical visits), two modalities (dNPI+clinical or biological+visits), then removing one of the 4 modalities. Here “medical records” stands for features extracted extracted from the medical record headers, that is to say, visit department and procedure. Performance scores are presented on the test set.

We also provide in the Supplementary Material a comparison of Tabular BEHRT with baselines that only make use of tabular information (Figure S6 in the Supplementary Material) and a comparison of Text BEHRT with baselines that only make use of text information (Figure S7 in the Supplementary Material). In both cases, the transformer-based approaches outperformed all comparison partners.

**Table 1.** AUC scores comparison for M-BEHRT and the baselines for the prediction of disease-free survival 3 years after surgery, on the test set. M-BEHRT significantly outperforms the other methods (DeLong test in Figure S22 in the Supplementaty Material).

| Models | AUC Scores |
| --- | --- |
| **M-BEHRT** | 0.77<br>[0.70 – 0.84] |
| NPI | 0.67<br>[0.58 – 0.76] |
| Random Forests | 0.67<br>[0.58 – 0.75] |
| Logistic Regression | 0.61<br>[0.52 – 0.71] |
| SVM | 0.61<br>[0.55 – 0.72] |

#### 3.2.3 Performance of M-BEHRT per cancer subtype

Figure 8 presents the AUC-ROC of M-BEHRT on the test set, stratified by patient age, tumor grade, molecular subtype, or node status. M-BEHRT is better at predicting DFS at three years on older patients, with at least one affected lymph node. Stratification of results by NPI range is available on Figure S8 in the Supplementary Material.

**Figure 8.**
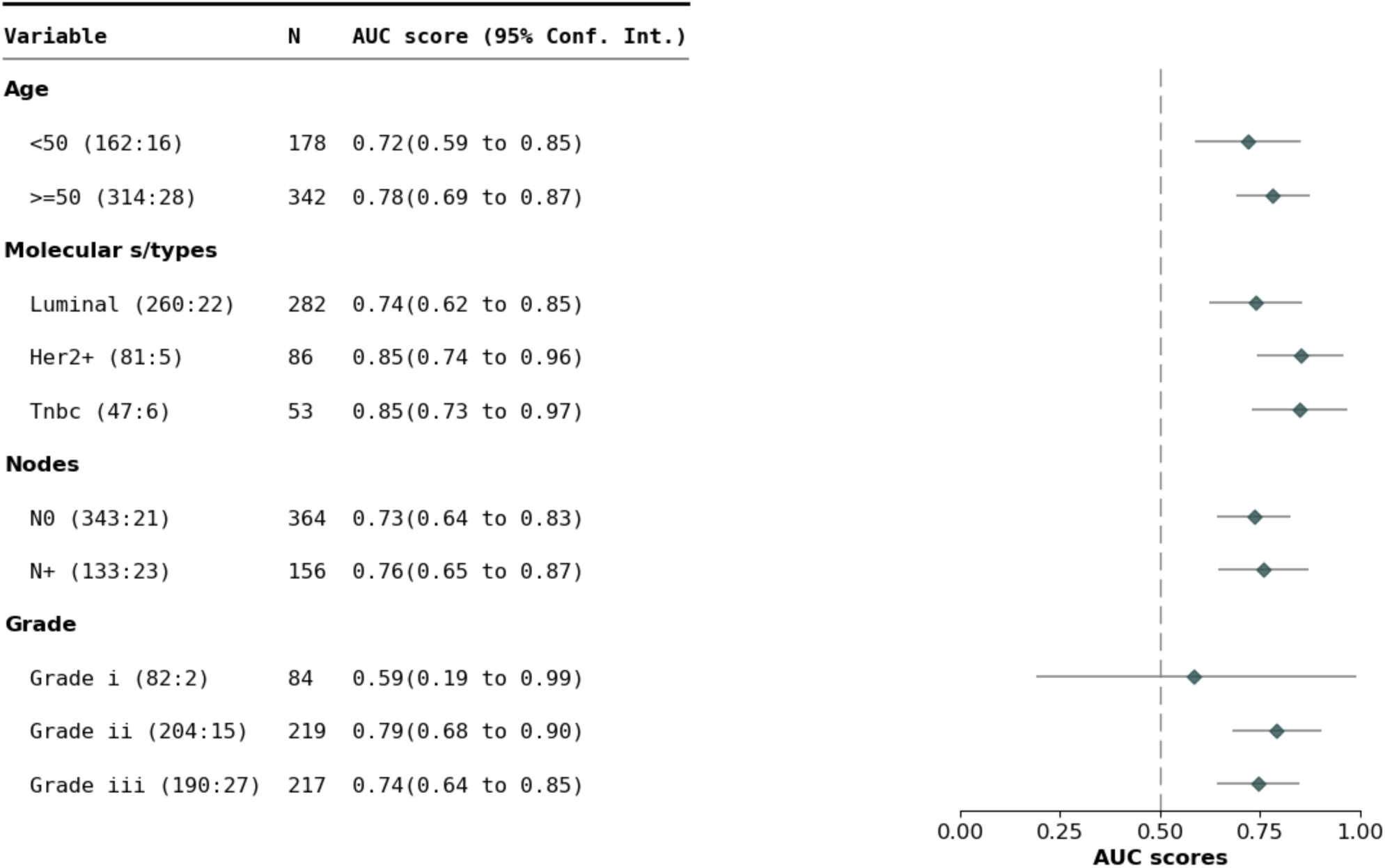
AUC-ROC of M-BEHRT on the test set stratified by patient age, cancer grade, molecular subtype and node status.

#### 3.2.4 Model interpretation

To better understand the predictions of M-BEHRT, we used the CAPTUM (32) implementation of the integrated gradients (IG) method (33) to attribute the predictions of either Tabular BEHRT or Text BEHRT to their input features. This allows us to highlight, for a given input sequence of visits, the elements that contributed to the label.

Overall, Tabular BEHRT mainly uses NPI tokens to correctly identify relapse or death for samples from the poor prognosis groups (VPPG and PPG), or to correctly identify DFS for patients from the good prognosis groups. What is more interesting, however, is to look at the tokens that Tabular BEHRT uses to accurately predict relapse or death for samples from the good and moderate prognosis groups, as they might provide critical insights into the aggressiveness and progression of the disease. They point towards having a high number of multidisciplinary consultation meetings (“RCP” in French), a high number of consultations overall, a second surgical procedure (within one year of the first one), or abnormal values for the CA15-3 and the LYMP biological markers. Moreover, Tabular BEHRT uses well-documented factors in the literature to predict a positive DFS status such as age.

The interpretation of Text BEHRT’s predictions shows that the model mostly relied on the entire sequence of the reports from the diagnosis to the index date to make its prediction, which is represented by the CLS token. We found this pattern in many true positive (correctly identifying death or relapse) samples. Moreover, Text BEHRT relies on reports that show information regarding the characterisation of a suspicious tumor, but this is not in and of itself indicative of a future relapse.

Finally, in order to gain a more global understanding of the model, we investigated the most predictive reports for a positive DFS status and for a negative DFS status. We set a threshold regarding the given attribution for each medical report. We collect all the reports with an attribution above this threshold. This yielded 921 reports that are predictive for negative DFS status in the entire corpus, and 1 720 reports that are predictive for positive DFS status. For each reports collection, we determined the 30 most frequent sequences (of 3 to 9 words) for both groups. We then listed the most frequent sequences for the DFS negative group that are not found in the DFS positive group. The resulting sequences of words can be found in Table 2.

**Table 2.** Most frequent sequences found in reports with high attribution for DFS-(relapse/death) instances but not for DFS+ instances, in Tabular BEHRT.

| Sequence meaning in English | Description |
| --- | --- |
| Breast in partial involution with less than 50% glandular tissue | Adipose involution is a natural process where glandular tissue is gradually replaced by fat tissue, often as a result of aging or hormonal changes. Here, the glandular tissue makes up less than half of the total breast composition. While age is a risk factor for breast cancer, lobular involution is associated with a reduced risk of breast cancer (34, 35). |
| Previous treatment with human growth hormone, without risk factors for CJD transmission | Treatment with human growth hormone can lead to the transmission of Creutzfeldt-Jakob disease (CJD). This information is a medical administrative criterion checked before surgery. |
| With axillary lymphadenectomy | Until recently, axillary lymph node dissection was standard procedure in the case of involvement of lymph node in breast cancer, one of the main known risk factors for relapse or death (36). |
| Palpable mass | Palpable breast lumps are the most common presentation of breast disease. |
| Solu-Medrol, 80mg | Solu-Medrol is one brand name for methylprednisolone, a corticosteroid used in BC to manage the side effects of taxane-based chemotherapy (37). |
| Lovenox 0.4 mL | Lovenox is one brand name for enoxaparin sodium, a low molecular weight heparin used as anticoagulant medication. It is used to prevent and treat venous thromboembolisms, for which cancer patients are at higher risk (38). |

Some of these sequences were obtained by combining overlapping sequences. We then plotted survival curves to compare patients that have reports containing one of these sentences and patients that do not. DFS is the event and the log-rank test is used to compare the populations. We show here two such curves, corresponding to sentences showing the most significant sequences: Figure 9 is for a sequence that translates to “breast in partial involution with less than 50% glandular tissue and Figure 10 is for a sequence that translates to “axillary lymphadenectomy”.

**Figure 9.**
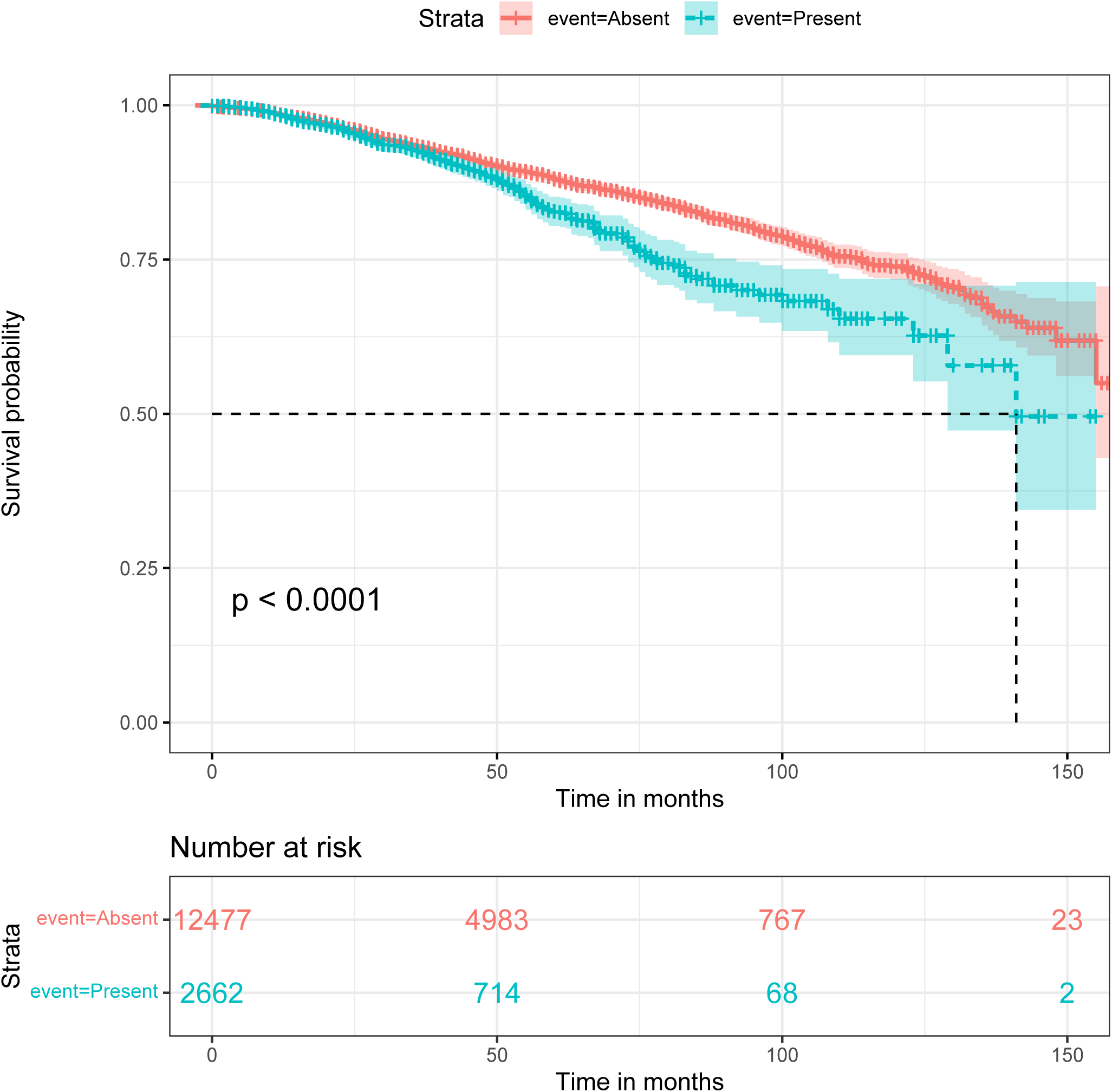
Survival plots for the sequence: “sein en involution adipeuse partielle avec contingent glandulaire inferieur a 50”, (*breast in partial involution with less than 50% glandular tissue*), Present or Absent in patients reports

**Figure 10.**
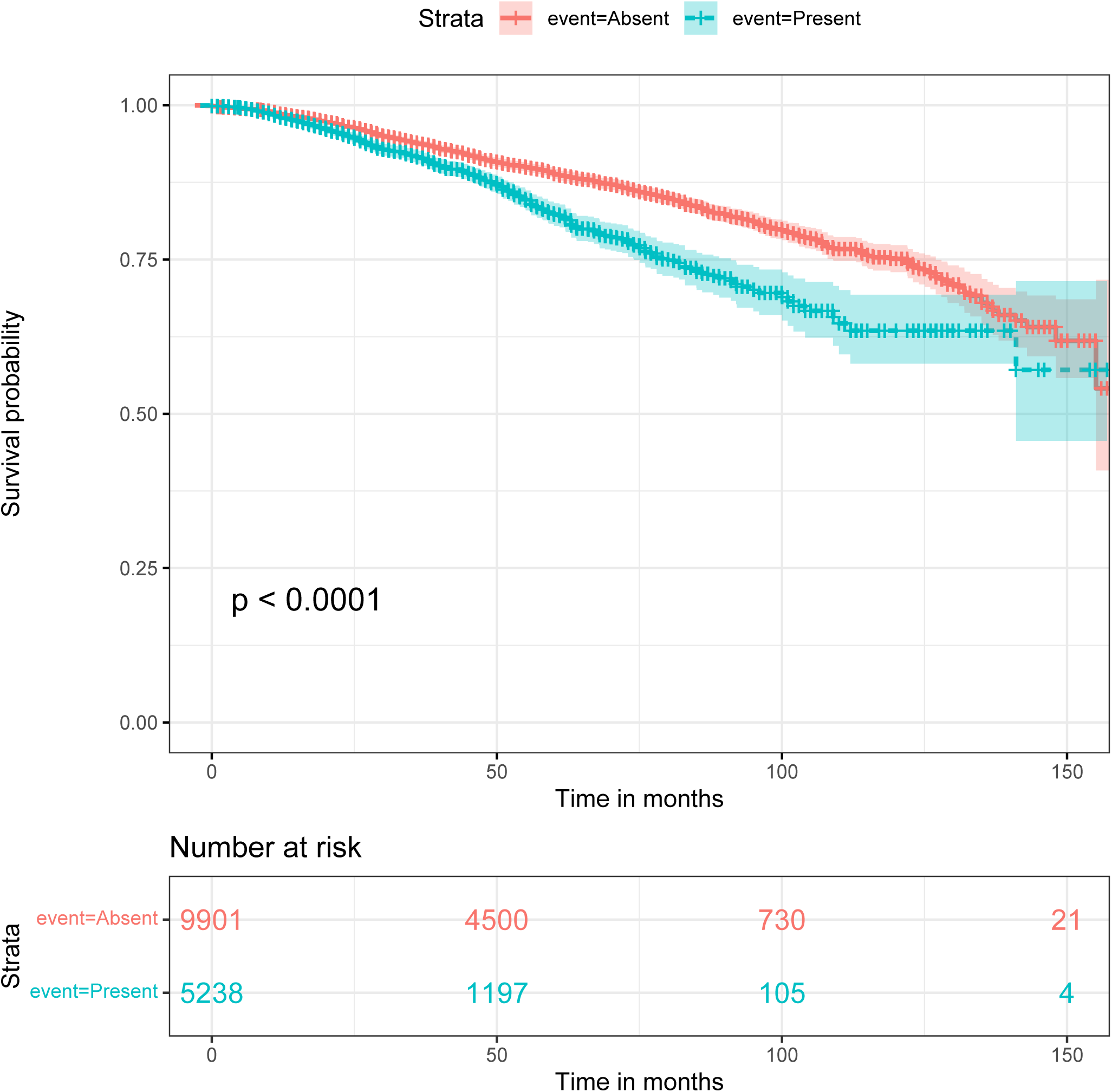
Survival plots for the sequence: “lymphadenectomie axillaire”, (*axillary lymphadenectomy*), Present or Absent in patients reports.

For the first example (Figure 9), the survival curves suggest that patients with this feature are most likely to relapse than others. This feature defines a specific state of breast tissue where the glandular tissue is replaced by adipose tissue. This process naturally occurs with aging and after menopause. Therefore, this feature could have an impact on DFS simply because it is related to the patient’s age, which is already a prognostic factor. However, when compared with 2 age groups (see Figure S11 in the Supplementary Materials), it added more information on the survival than just *>* 50 years old and *<* 50 years old. Young patients with this feature represent the worst prognostic groups.

Although mammary involution is not a commonly used prognostic factor, several studies have showed a link between involution and breast cancer risk (34, 35); the underlying biological process could maybe also explain a hightened risk of relapse in young patients presenting abnormal mammary involution.

The second plot (Figure 10) compared a population with the feature “axillary lymphadenectomy” and a population without. This feature is a mention of removing lymph nodes from the armpits. This information is associated with the potential affection of axillary nodes, which is found to be predictive for BC relapse.

## 4 DISCUSSION

In this paper, we proposed several novel deep learning architectures inspired by BEHRT to model patient trajectories using multimodal data extracted from EHRs. As the original BEHRT model, Tabular BEHRT considers structured data to describe each medical event. In addition, it considers multiple modalities (biological lab results, clinical information, department and procedure names) simultaneously. By contrast, in Text BEHRT each visit is described via the content of free text medical reports. Finally, M-BEHRT combines both models through cross-attention. Our work is motivated by applications to oncology, and applied to the prediction of disease-free survival for breast cancer patients.

### 4.1 M-BEHRT achieves state-of-the-art or better prediction of DFS

Using very different information, Tabular BEHRT and Text BEHRT achieve AUCs on a held-out data set of 0.75 [0.66-0.83] and 0.75 [0.68-0.81], respectively, for the prediction of DFS 3 years after surgery. Combining them in M-BEHRT slightly increases predictive power, reaching an AUC of 0.77 [0.70-0.84]. All three architectures outperform classical machine learning methods. M-BEHRT is therefore able to capture the sequential aspect of patient data throughout their medical journey, resulting in improved performance.

To date, most of the multimodal prognosis models for breast cancer use various types of medical images, as well as sometimes genetics data, combined or not with tabular information (biological measurements, clinical features). Moreover, endpoints vary between studies: DFS, but also overall survival or recurrence (sometimes separated between local, regional and distant); which can be measured 3 years after surgery as in the present work, but also at different time points. Finally, different studies use different criteria inclusions. All in all, this makes comparing our performance to other studies challenging. However, we note that M-BEHRT achieves better performance for the prediction of DFS after three years than the recent work of Han et al. (6), which uses ultrasound and mammography images combined with clinical, pathological and radiographic characteristics and reports an AUC of 0.739 on a held-out test set. In addition, the performance of M-BEHRT is in the same ballpark as that of Rabinovici-Cohen et al. (5), which predict recurrence at five years in patients who receive neo-adjuvent chemotherapy (AUC of 0.75 on a held out data set) using clinical features, immunohistochemical markers, and multiparametric magnetic resonance imaging, or González-Castro et al. (9), which achieve an AUC of 0.81 also for predicting recurrence at five years, but considering all cancer patients and using clinical features, immunohistochemical markers, and descriptors of clinical history such as the number and type of therapies.

In order to further evaluate the ability of M-BEHRT to predict DFS, we also performed the same study, but for the prediction of DFS 5 years after surgery rather than 3. This results in a smaller data set of 5 192 patients. The test set is the same as for DFS 3 years after surgery, but now contains 17.1% of negative samples. All results are available in the Supplementary Materials (Table S4 and Figure S10 for a description of the data, and Figures S11-17 for the results). Our observations are similar to those made on the prediction of DFS 3 years after surgery, although predicting DFS 3 years after surgery seems much easier than 5 years after surgery (AUC of 0.77 vs 0.69). This is in line with previous observations that earlier events are easier to predict than long-term ones (39).

We stratified the data based on features that are expected to define patients with similar prognoses (age, grade, number of lymph nodes involved, molecular subtype). We found that the prediction ability of M-BEHRT varies depending on subgroups and that the model works better on older patients with more aggressive disease (at least one lymph node involved). In addition, M-BEHRT is better at predicting relapse after 5 years than after 3 years for luminal tumors, suggesting that it correctly identifies predictive factors with long term influence for these tumors that tend to recur later than others (40).

There are however some limitations to the scope of our study. In particular, our findings are restricted to a very specific cohort of patients who received adjuvant chemotherapy. We also have not been able to validate our findings on an external validation group, due to privacy concerns limiting the access to EHR of other centers; it is possible that our models have captured idiosyncrasies of Institut Curie that do not apply to patients from other hospitals. However, our work shows that it is possible to learn from multimodal patient trajectories built from dynamic tabular data and the content of free-text reports written by practicioners at each medical visit, and paves the way for future research in understanding breast cancer prognostic factors.

### 4.2 M-BEHRT learns on small data sets

An important aspect of our study is that, unlike most work published to date using transformers for EHR data, which use millions of patients for pretraining and tens to hundreds of thousands of patients for fine-tuning (11, 13, 12), the datasets we use here are of much smaller sizes: about 15 000 patients for pretraining, and 5 000 to 8 000 patients for fine-tuning. That it is possible to apply such methods to much smaller data sets is very encouraging for future research, as many studies, especially on very specific diseases and endpoints, only have access to a limited number of patients.

However, despite the small sample size, our study has an advantage over those with larger datasets’ studies because our learning data includes only adjuvant-treated breast cancer patients. This specificity has enabled the model to learn more precise embeddings and improve the accuracy of relapse prediction.

Keeping the same pretrained model, we experimented with further reducing the number of patients used for training the classifier. To this end, we created smaller training sets by randomly selecting subsets of the training data, starting from 10 samples, and compared on the test set the performance of Tabular BEHRT and classical machine learning algorithms trained on these small training sets. Our results, shown on Figure S20 in the Supplementary Material, show that Tabular BEHRT clearly outperforms the classical machine learning algorithms, especially random forests, in the few-shot learning setting (when training set sizes are very small), achieving better-than-random performance with as little as 10 training samples and outperforming NPI with a few hundred training samples. We attribute this performance to the ability of the pretraining phase to learn meaningful representations of patient trajectories.

### 4.3 M-BEHRT leverages the complementary nature of different modalities

In order to better understand the contribution of the different modalities to the performance of Tabular BEHRT, we conducted an ablation study. The results show that, with the exception of the biological features, excluding one modality or more substantially reduces model performance. This indicates that Tabular BEHRT has the ability to leverage the complementary nature of the different modalities. In addition, clinical features (dNPI, age, molecular subtype and therapy) contribute the most to performance. This observation is consistent with previous studies on breast cancer relapse prediction (41, 42).

Although others have found the results of routine laboratory tests to be very informative for predicting breast cancer endpoints (4, 41), our study did not see strong added value of including biological markers on DFS prediction. This is particularly surprising regarding cancer antigen CA 15-3, which has been found in several stiduies to correlate to poor prognosis (43, 44) and recurrence (45, 41). In addition, Kim et al. (41) found that an increase in leukocyte count (LEUK) has a protective effect against breast cancer recurrence and that an elevated neutrophil count (PN) is associated with recurrence, although another study (4) did not find a significant association between DFS and variables describing leukocyte counts and counts or percentages of leukocyte subtypes. However, these features not entirely uninformative, as restricting Tabular BEHRT to the biological features modality still yields better-than-random performance (AUC of 0.56 for T1 and 0.61 for T2). One possible explanation is that the information contained in the biological features is also captured by the other modalities, as their evolution might be consistent with cancer severity or subtype, or the choice of therapy. Our study is also limited in the number of available laboratory variables, as markers that were found informative in previous studies, such as hemoglobin, total protein, serum glucose, alkaline phosphatase, or international normalized ratio (41, 4) were not available (or not for enough patients) in our data.

Perhaps surprisingly, we do not see the same drastic increase in performance between Tabular BEHRT and M-BEHRT as others have observed in multimodal prediction of breast cancer prognosis when augmenting clinical data with imaging data (5, 6), although Text BEHRT leverages medical reports from radiologists or cytopathologists, which are based on medical images. Although this could be due to the aforementioned limitations of Text BEHRT, this could also be because Tabular BEHRT already achieves much better performance than models based solely on static clinical data.

### 4.4 M-BEHRT model interpretation points to possible prognostic factors

The interpretation of M-BEHRT models through the integrated gradients method highlighted that Tabular BEHRT relies on well-documented prognostic features such as the age or the NPI (46, 2) to predict DFS status. Additionnally, the model uses features that indicate a more aggressive breast cancer (number of multidisciplinary meetings, number of consultations, or a second surgical procedure), which can not be necessarily be considered as causes of cancer relapse but suggest a more difficult-to-treat cancer.

Regarding Text BEHRT, the model seems to rely mainly on reports that contain symptoms-related information or reports from imagery. When they occur before the first surgery, these information are to be expected, as we are studying a cohort of patients treated for breast cancer. However, if they occur after the first surgery, these features can indicate further investigations that are warranted by the difficulty to treat the primary tumor.

Let us note however that while deep learning model interpretation is still somewhat limited, it has the potential to offer a much more comprehensive interpretation of the roles played by different elements in the data, given how rich the data is. Moreover, the features that are highlighted as strongly contributing towards one label or the other are only doing so in conjunction with other features, which might be different from patient to patient. Moreover, the embedding pooling method that we have used to derive reports embeddings from their contents does not help with interpretability, as it does not allow to pinpoint specific parts of a medical report. Nevertheless, several potentially interesting text features (such as high mammary involution or axillary dissection) have been highlighted for their contribution to M-BEHRT predictions. Even though it is not yet clear how these features can be used as prognostic factors and incorporated in a model usable in the clinic, survival curves show that they are indeed informative of DFS even taken on their own.

### 4.5 Challenges of learning from long sequences of rich events

In our approach, there is a tradeoff between the number of visits that can be considered and the amount of information that can be used to describe each visit, because the underlying BERT architecture is limited to processing 512 tokens. This number is arbitrary, but constrained by the memory usage of the self-attention mechanism. We have found this number to be sufficient for the DFS prediction tasks at hand and the available features and modalities. However, this might be too small for other applications, in which case one might want to use approaches that approximate the self-attention matrices so as to reduce their memory footprint, such as Big Bird (22) or Nyströmformer (23). In the present study, M-BEHRT outperforms both NPI and classical ML baselines, suggesting its ability to capture the structure of EHR data.

To the best of our knowledge, ours is the first study to use entire free text medical reports (in a language other than English) for breast cancer prognosis. There are several limitations to our approach. First, we used token embeddings learned on French clinical text that are not specific to breast cancer; it is possible that pretraining on breast cancer clinical text could improve the performance of our model. However, this requires considerable resources, both in terms of amount of clinical records available and computing power. Second, we build medical records embedding by simply pooling all token embeddings of a record, which is likely not be optimal for capturing the information contained in a report. Several authors have proposed using convolutional neural networks (CNN) or bidirectional long-short term memory architectures (Bi-LSTM) on top of token embeddings (20, 47, 48), which typically helps capturing the structure of text documents and could be an interesting future direction to explore for this research. Despite these shortcomings, our results demonstrate the ability of Text BEHRT to capture relevant information, as it performs on par with Tabular BEHRT.

Finally, M-BEHRT uses a cross-attention module to perform the multimodal fusion between Tabular BEHRT and Text BEHRT. This approach allows the contextual integration of information from both transformers, i.e, that each model can attend information from the other model, and thus enable a better exploitation of the complementarity between inputs. However, this requires that both tabular data and text data embeddings have the same size, and forced us to reduce the dimensionality of the embedding of sequences of reportsfrom 768 (as provided by DrBERT) to 144 through a linear layer. This may result in an additional reduction of available information. However, this still results in a slight improvement of overall performance.

### 4.6 Conclusion

Overall, our study highlights the potential to predict DFS using solely longitudinal sequence of medical visits and evolution of clinical information and biological measurements. To the best of our knowledge, this is the first study predicting breast cancer endpoints from sequences of EHR data, whether considering solely multimodal dynamic tabular data, solely the contents of free-text reports, or combining both. Our results underscore the usefulness of such data for future research on prognosis modeling, and outline the importance of integrating medical information collected over time to gain previously unknown insights into the understanding of breast cancer evolution.

## Data Availability

Data can only be accessed once approval has been obtained through the Institutional Review Board of Institut Curie.

## ETHICS STATEMENT

The studies involving human participants were reviewed and approved by the Institutional Review Board of Institut Curie (Paris, France). The patients/participants provided their written informed consent to participate in this study.

## AUTHOR CONTRIBUTIONS

Conceptualization: CAA, MD, MRZ, NMB

Data curation: JG, ED, AHP, AT

Formal analysis: NMB

Funding acquisition: CAA, MRZ, FR

Investigation: NMB, AT

Methodology: CAA, MD, MRZ, NMB

Project administration: CAA, NMB

Resources: JG

Software: NMB

Supervision: CAA, MRZ

Visualization: NMB

Writing – original draft: CAA, NMB

Writing – review & editing: CAA, NMB, MRZ, MD

## ACKNOWLEDGMENTS

The authors thank Éric Daoud, Antoine Recanati and Charles Vesteghem for fruitful discussion and Johan Archinard for the technical environment maintenance.

## DATA AVAILABILITY STATEMENT

Electronic health records are considered sensitive data in the EU by the General Data Protection Regulation and cannot be shared via public deposition because of legal restriction in place to protect patient confidentiality. Data can only be accessed once approval has been obtained through the Institutional Review Board of Institut Curie.

## Supplementary Material to Multimodal BEHRT: Transformers for Multimodal Electronical Health Records

### 1 SUPPLEMENTARY TABLES AND FIGURES

**Table S1.** Normal ranges for the biological features.

| Feature | Normal range | Mean value ± std | missing |
| --- | --- | --- | --- |
| CA15-3 (U/ml) | $N < 30$ | $63.39 \pm 484.44$ | 6 390 |
| LEUK (g/l) | $4 < N < 10$ | $6.99 \pm 6.82$ | 2 525 |
| PN (g/l) | $1.7 < N < 7$ | $718.85 \pm 1\,789.66$ | 9 419 |
| LYMP (g/l) | $1.4 < N < 4$ | $289.63 \pm 714.26$ | 9 448 |
| MONO (g/l) | $0.2 < N < 1$ | $33.29 \pm 123.59$ | 3 675 |

**Table S2.** Descriptive statistics of the data used in this study, for the full cohort of 15 150 patients, as well as the data set of patients uncensored 3 years after surgery.

| Features |  | Entire dataset |  | Dataset for DFS at 3 years |  |
| --- | --- | --- | --- | --- | --- |
| | | Mean ± std | N | Mean ± std | N |
| Age | $< 50$ | $58 \pm 12$ | 3 982 | $56 \pm 12$ | 2 493 |
| | $\geq 50$ | | 11 168 | | 5 596 |
| BC subtype | Luminal |  | 9 979 |  | 4 866 |
|  | TNBC |  | 1 041 |  | 642 |
|  | HER2+/HR+ |  | 681 |  | 587 |
|  | HER2+/HR- |  | 480 |  | 415 |
| Grades | I |  | 3 473 |  | 1 688 |
|  | II |  | 5 911 |  | 3 057 |
|  | III |  | 3 119 |  | 2 044 |
| Nodes | N0 | $0.93 \pm 2.49$ | 9 463 | $1.07 \pm 2.74$ | 4 899 |
|  | N+ |  | 4 045 |  | 2 405 |
| Tumor size (mm) | Clinical | $16.89 \pm 12.70$ | | $17.36 \pm 12.97$ | |
| | Pathological | $15.04 \pm 12.75$ | | $15.63 \pm 12.90$ | |
| Biological values | CA 15-3 (U/ml) | $63.39 \pm 484.44$ | 8 760 | $62.85 \pm 535.76$ | 3 826 |
| | LEUK (g/l) | $6.99 \pm 6.82$ | 12 625 | $6.90 \pm 7.49$ | 6 419 |
| | PN (g/l) | $718.85 \pm 1\,789.66$ | 5 731 | $976.17 \pm 2\,007.52$ | 2 385 |
| | LYMP (g/l) | $289.63 \pm 714.26$ | 5 702 | $405.84 \pm 820.08$ | 2 373 |
| | MONO (g/l) | $33.29 \pm 123.59$ | 11 475 | $37.54 \pm 131.79$ | 5 821 |
| Medical reports | visits | $46 \pm 33$ | | $25 \pm 10$ | |
| | reports | $62 \pm 50$ | | $34 \pm 15$ | |
| | words/report | $172 \pm 41$ | | $159 \pm 37$ | |

**Figure S1.**
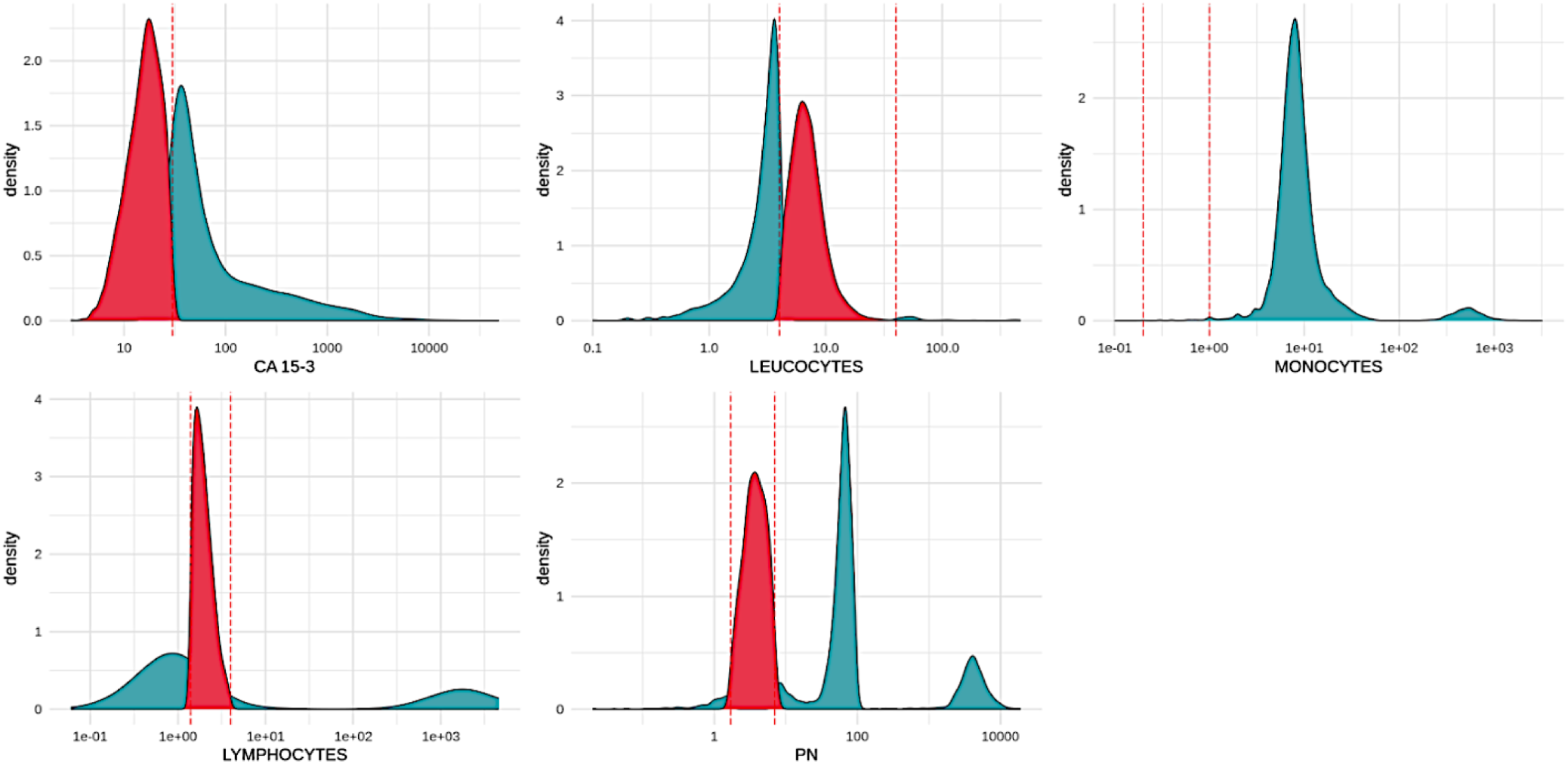
Binarization of biological features into two values, 1 and 2. For each of the 5 biological features, the dashed red lines delineate the normal range, highlighted in red, and mapped to 2, from the abnormal range, highlighted in green, and mapped to 1

**Table S3.**
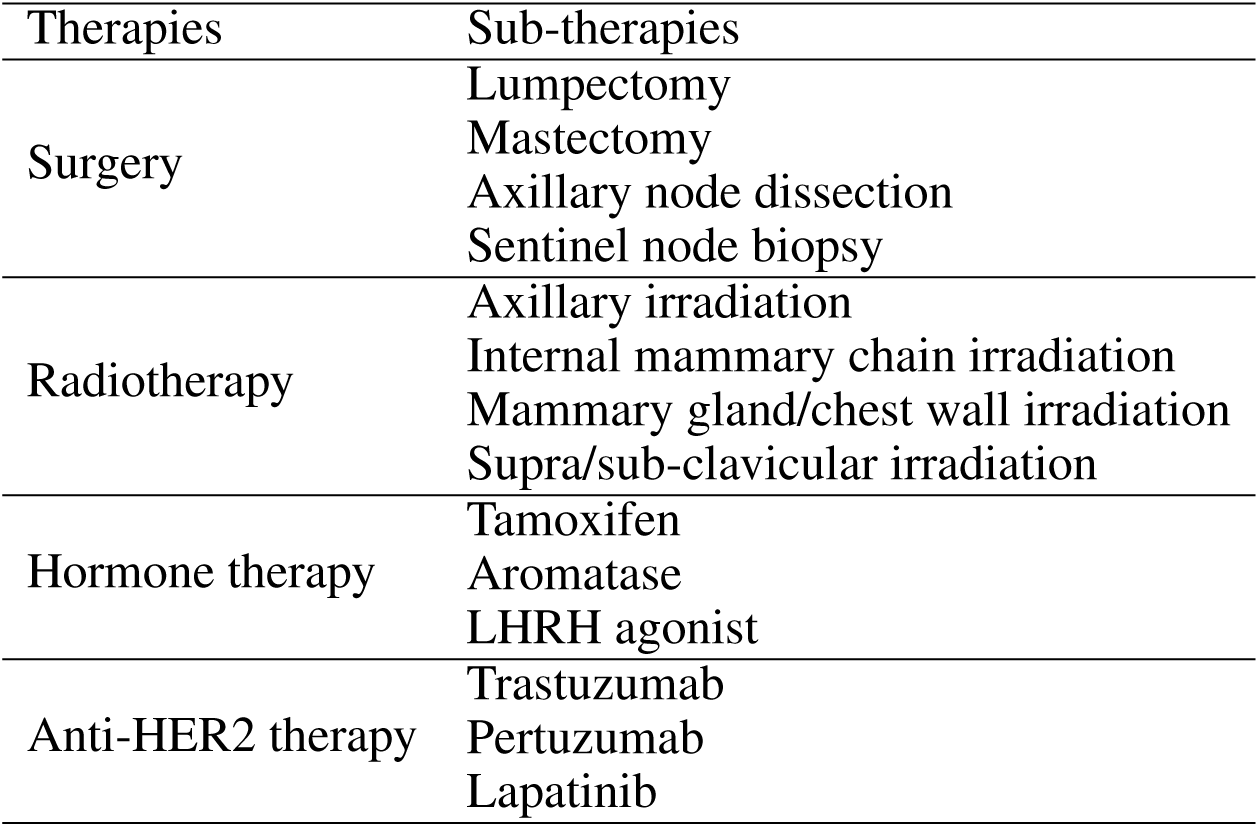
List of possible therapies and sub-therapies in our data.

**Figure S2.**
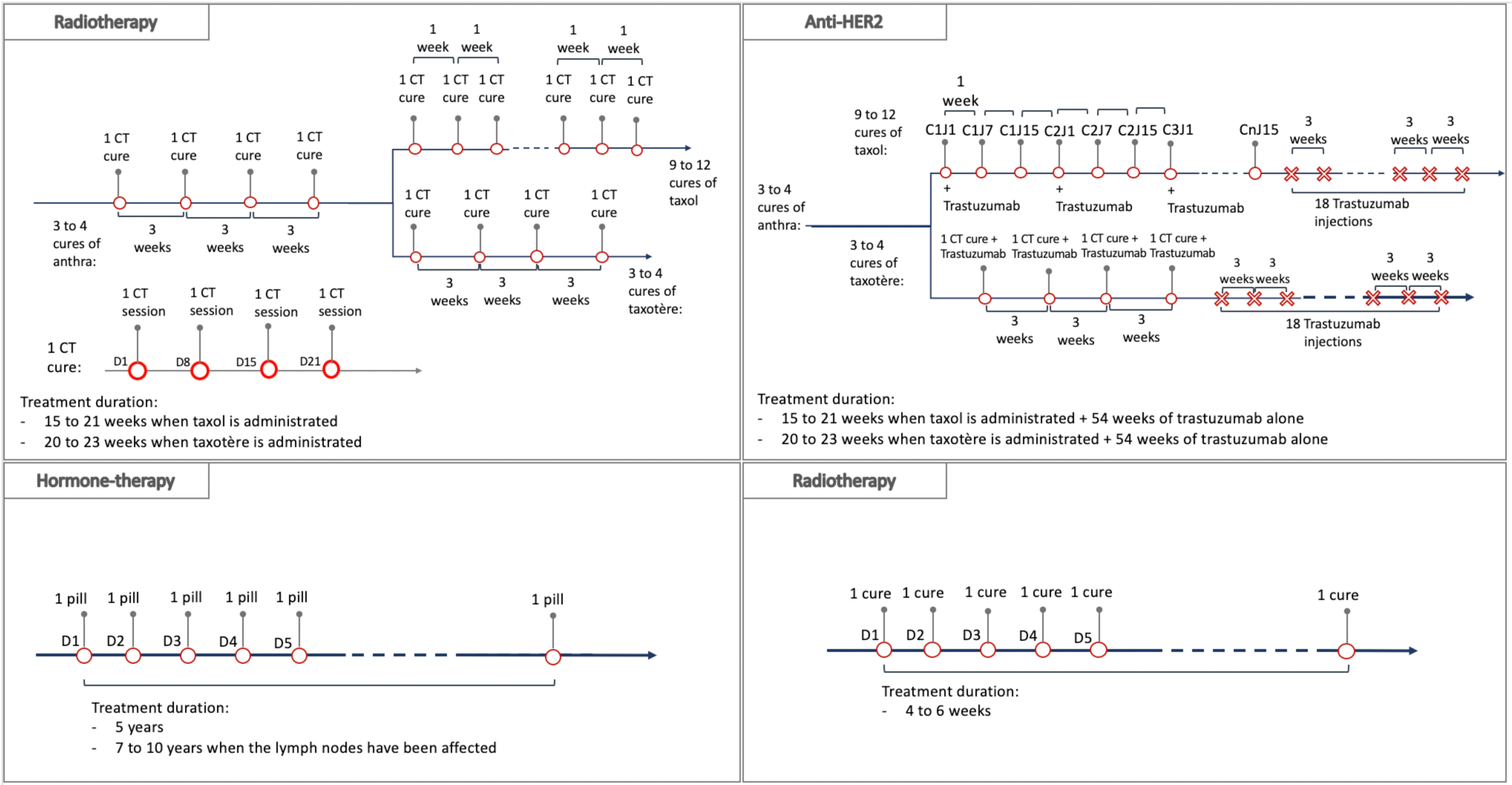
Institut Curie Therapeutic Protocol.

**Figure S3.**
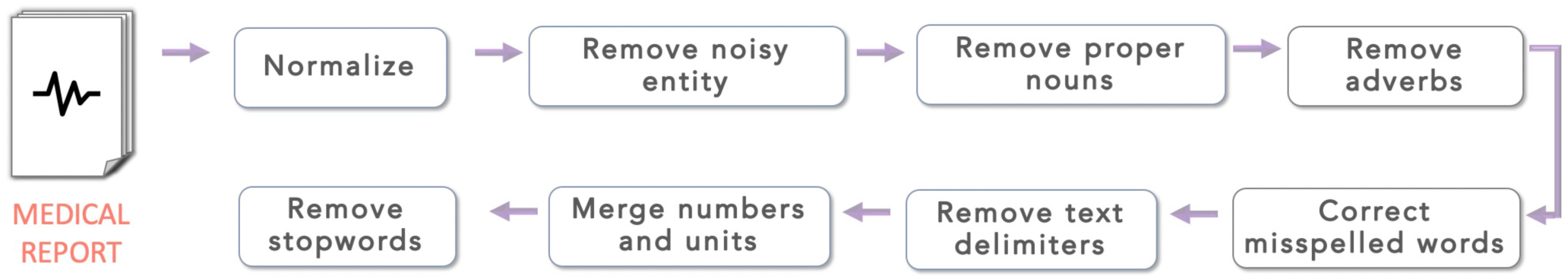
Text preprocessing pipeline.

**Figure S4.**
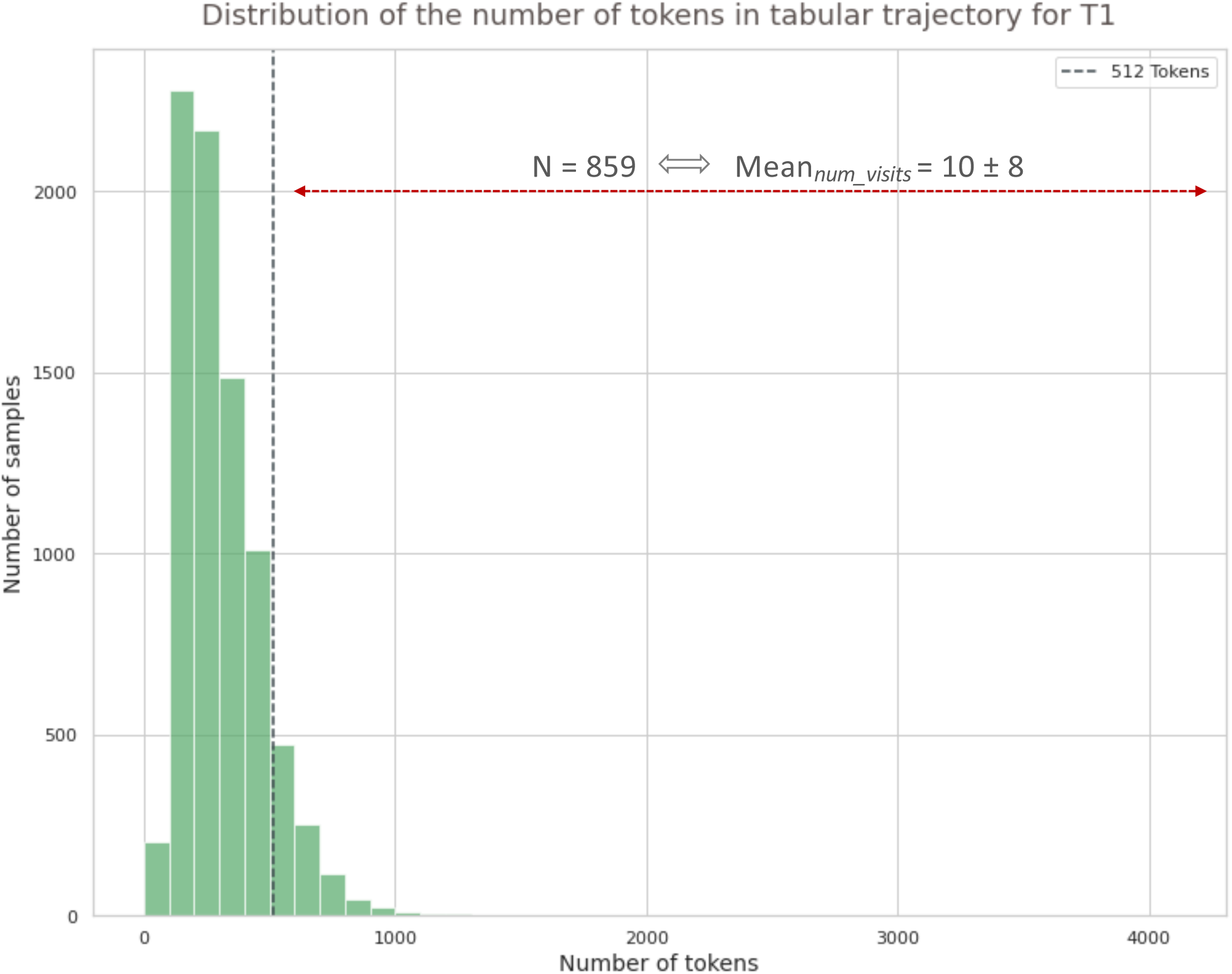
Distribution of the number of tokens per patient trajectory, for the prediction of disease-free survival 3 years after surgery. 859 samples exceed the maximum sequence length for Tabular BEHRT (512 tokens). This represents an average of 10 visits per patient that are not considered by Tabular BEHRT.

**Figure S5.**
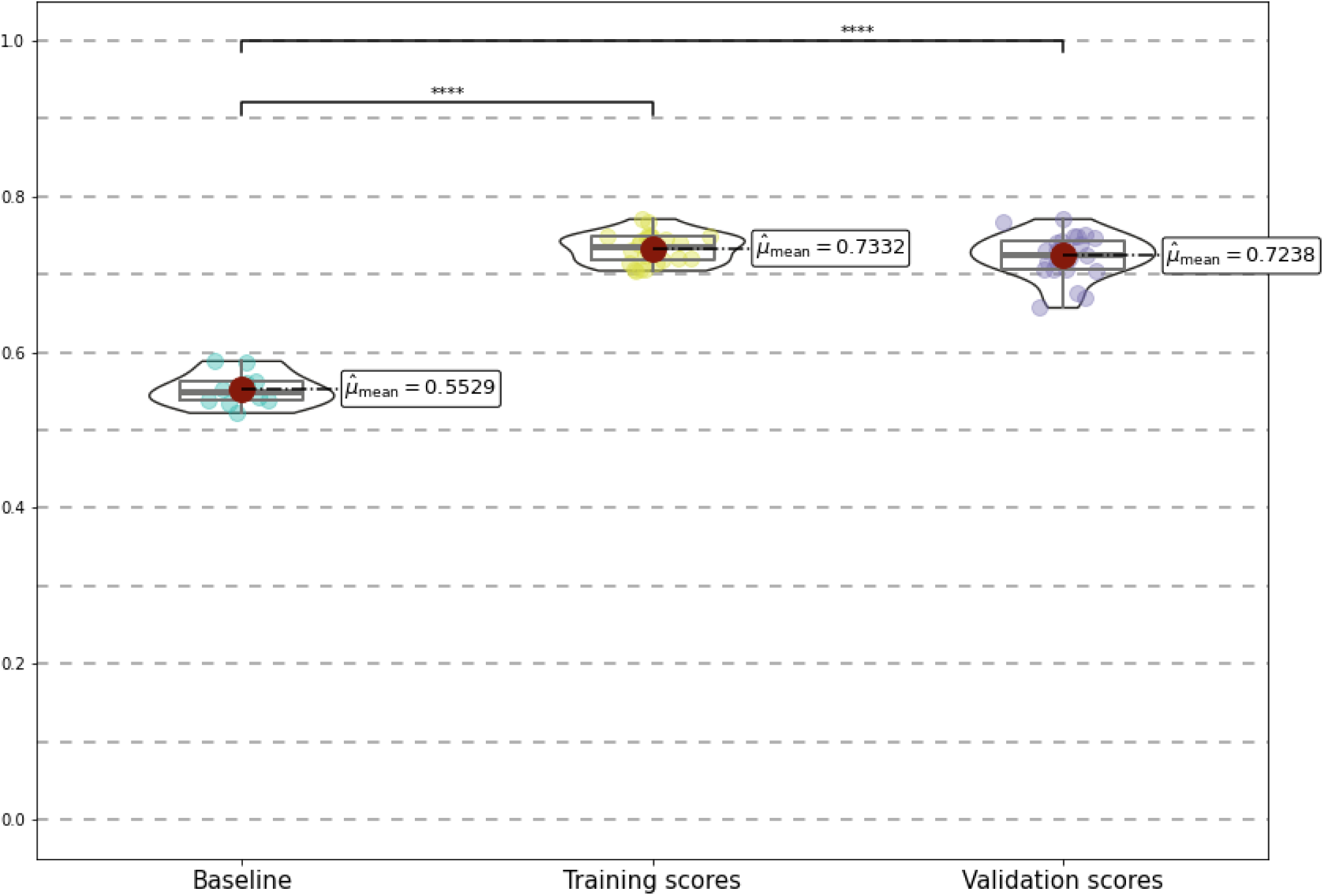
Precision scores for the Masked Language Model (pre-training of Tabular BEHRT). The baseline scores are obtained from the MLM ran on shuffled sequences.

**Figure S6.**
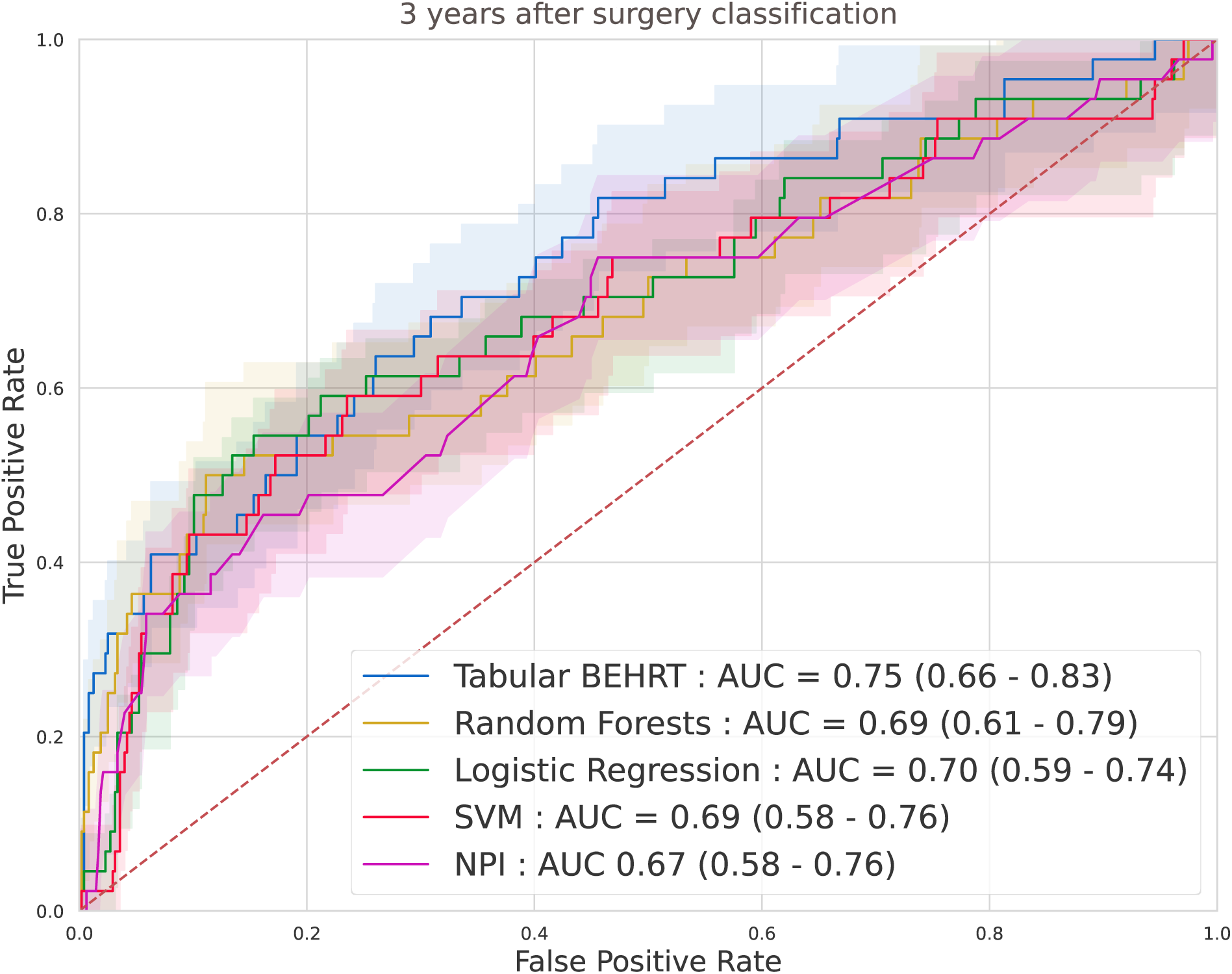
ROC curves for baselines and Tabular BEHRT, for predicting disease-free survival 3 years after surgery.

**Figure S7.**
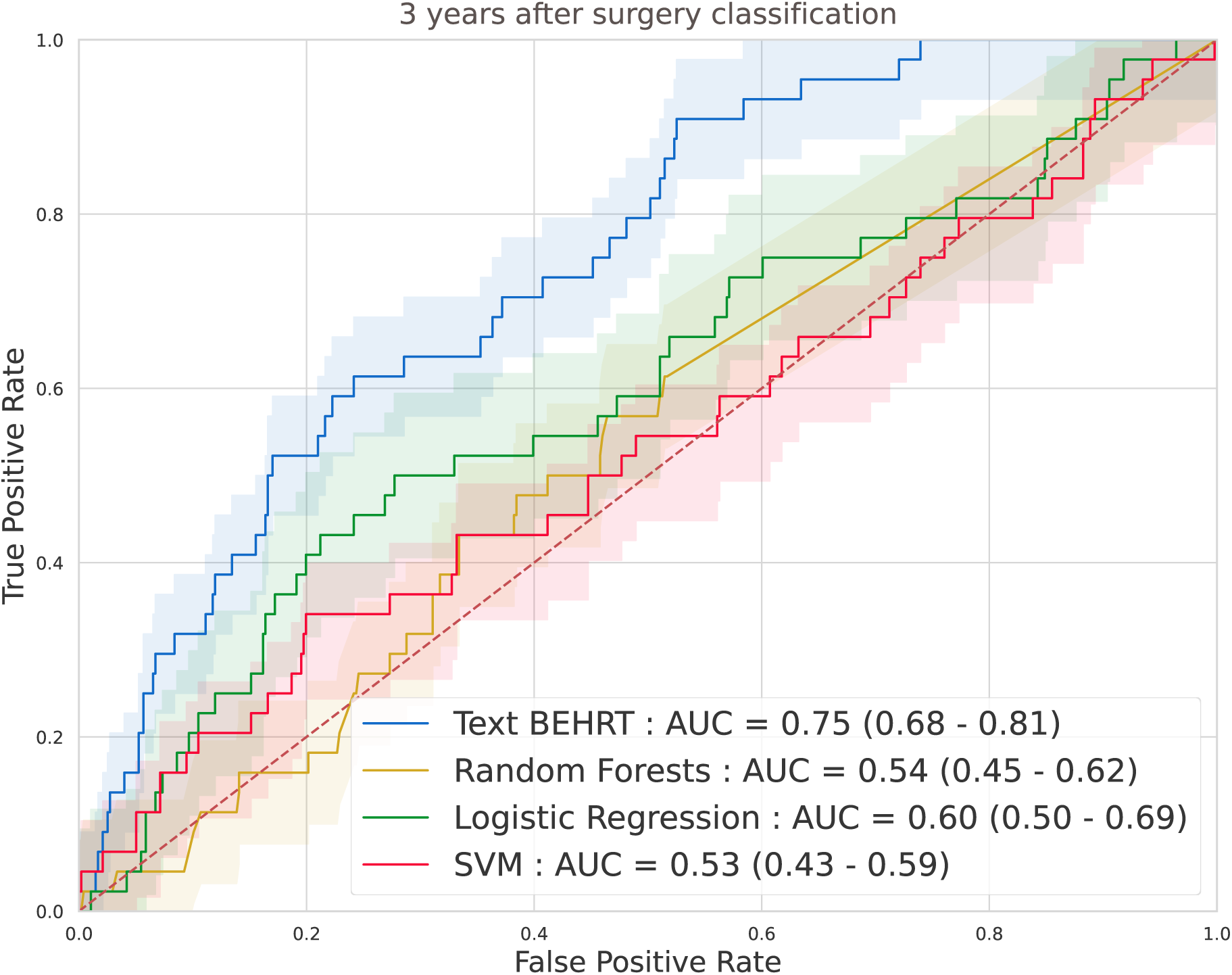
ROC curves for baselines and Text BEHRT, for predicting disease-free survival 3 years after surgery.

**Figure S8.**
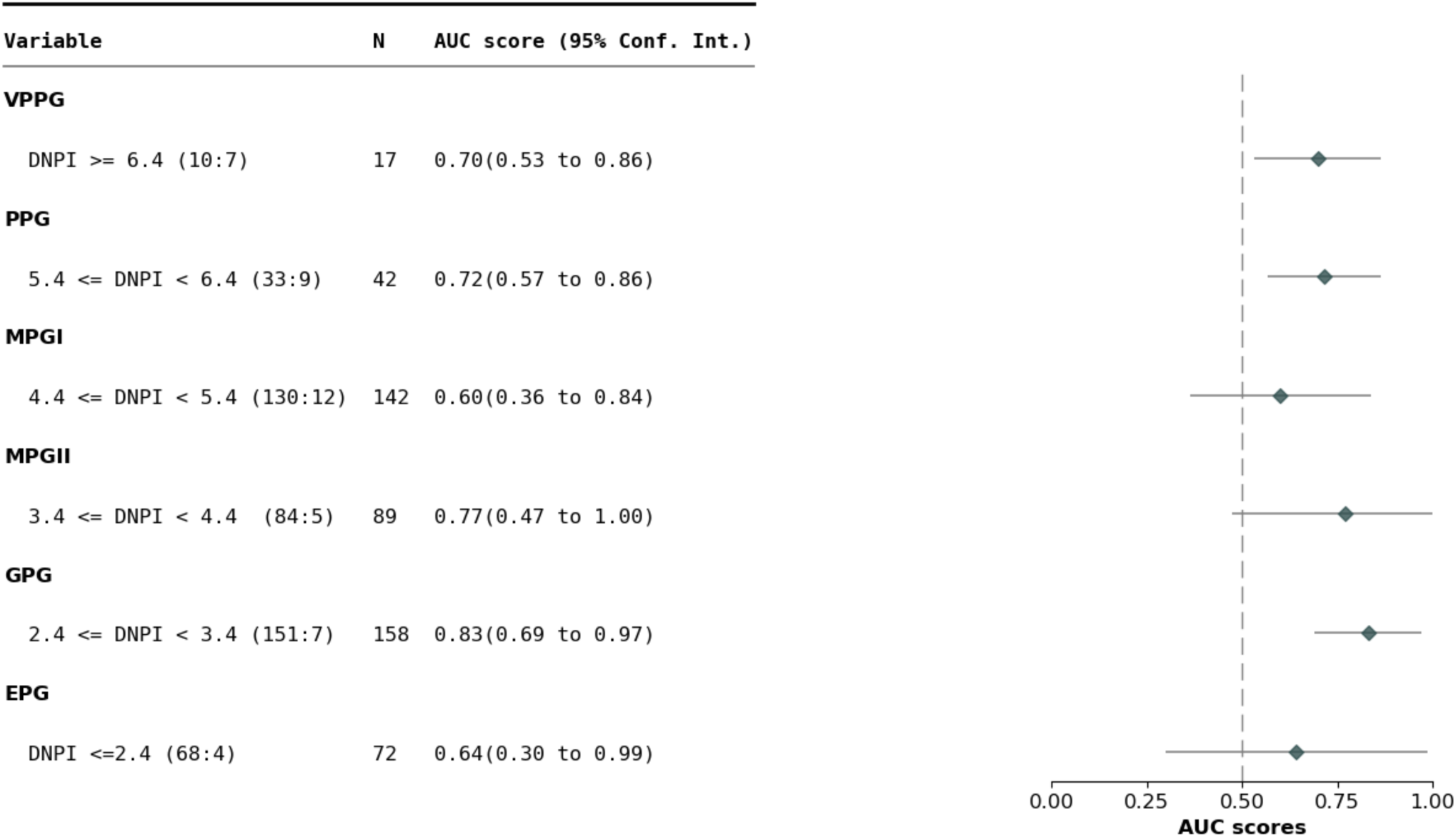
AUC-ROC of M-BEHRT stratified by NPI, for predicting disease-free survival 3 years after surgery.

**Table S4.** Descriptive statistics of the data sets used in this study, for the full cohort of 15 150 patients, as well as the data set of patients uncensored 3 years and 5 years after surgery.

| Features |  | Entire dataset |  | Dataset for DFS at 3 years |  | Dataset for DFS at 5 years |  |
| --- | --- | --- | --- | --- | --- | --- | --- |
| | | Mean ± std | N | Mean ± std | N | Mean ± std | N |
| Age | < 50 | 58 ± 12 | 3 982 | 56 ± 12 | 2 493 | 55 ± 13 | 1 725 |
| | $\geq 50$ | | 11 168 | | 5 596 | | 3 467 |
| BC subtype | Luminal |  | 9 979 |  | 4 866 |  | 2 930 |
|  | TNBC |  | 1 041 |  | 642 |  | 446 |
|  | HER2+/HR+ |  | 681 |  | 587 |  | 482 |
|  | HER2+/HR- |  | 480 |  | 415 |  | 330 |
| Grades | I |  | 3 473 |  | 1 688 |  | 1 016 |
|  | II |  | 5 911 |  | 3 057 |  | 1 941 |
|  | III |  | 3 119 |  | 2 044 |  | 1 462 |
| Nodes | N0 | 0.93 ± 2.49 | 9 463 | 1.07 ± 2.74 | 4 899 | 1.18 ± 3.01 | 3 132 |
|  | N+ |  | 4 045 |  | 2 405 |  | 1 597 |
| Tumor size (mm) | Clinical | 16.89 ± 12.70 | | 17.36 ± 12.97 | | 17.78 ± 13.18 | |
| | Pathological | 15.04 ± 12.75 | | 15.63 ± 12.90 | | 16.17 ± 12.94 | |
| Biological values | CA 15-3 (U/ml) | 63.39 ± 484.44 | 8 760 | 62.85 ± 535.76 | 3 826 | 75.34 ± 617.09 | 2 256 |
| | LEUK (g/l) | 6.99 ± 6.82 | 12 625 | 6.90 ± 7.49 | 6 419 | 6.75 ± 3.60 | 3 916 |
| | PN (g/l) | 718.85 ± 1789.66 | 5 731 | 976.17 ± 2007.52 | 2 385 | 1105.76 ± 2093.81 | 1 365 |
| | LYMP (g/l) | 289.63 ± 714.26 | 5 702 | 405.84 ± 820.08 | 2 373 | 463.92 ± 862.37 | 1 375 |
| | MONO (g/l) | 33.29 ± 123.59 | 11 475 | 37.54 ± 131.79 | 5 821 | 33.29 ± 123.59 | 3 489 |
| Medical reports | visits | 46 ± 33 | | 25 ± 10 | | 25 ± 10 | |
| | reports | 62 ± 50 | | 34 ± 15 | | 34 ± 15 | |
| | words/report | 172 ± 41 | | 159 ± 37 | | 159 ± 37 | |

**Figure S9.**
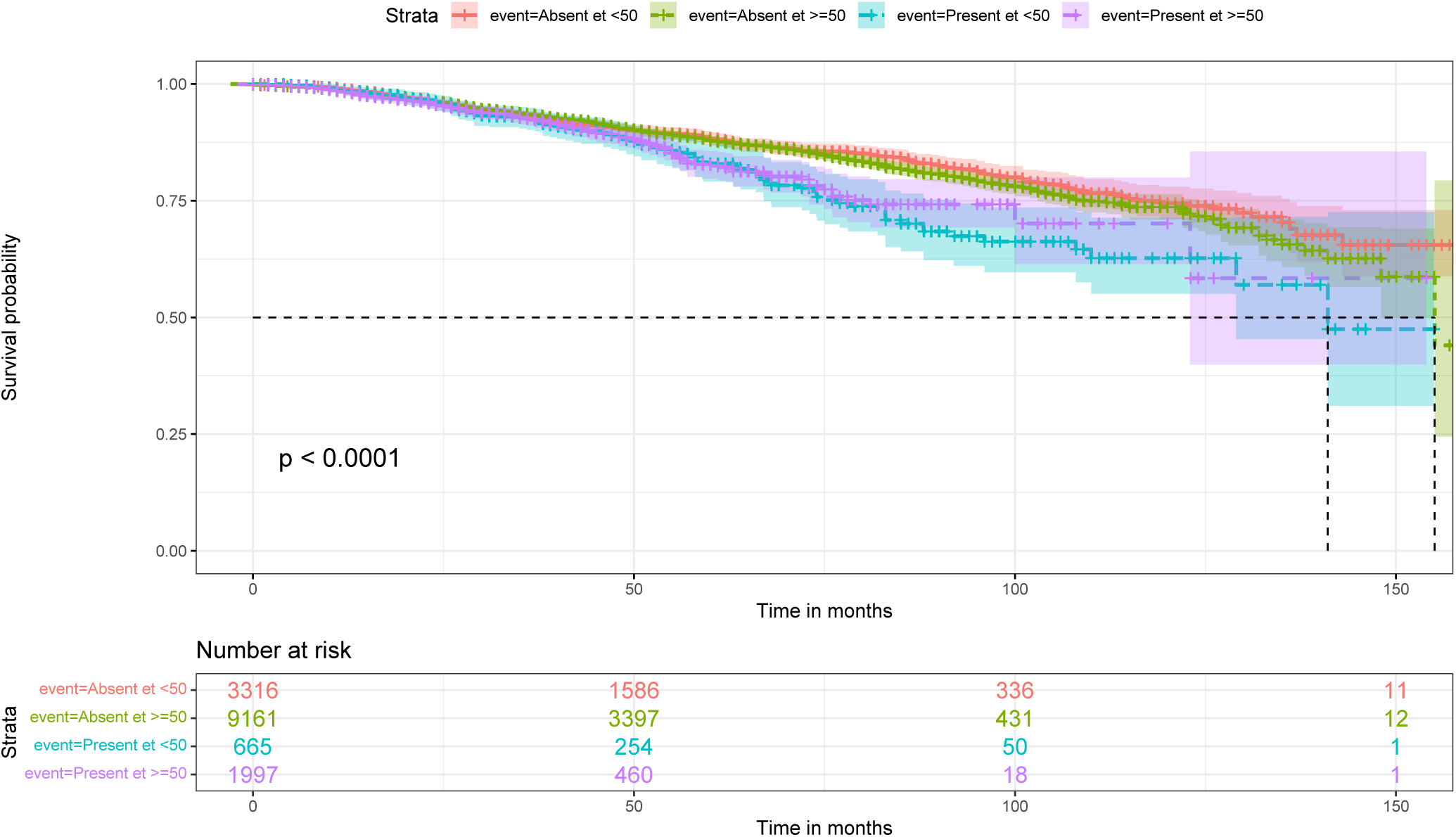
Survival plots for the presence/absence of the sentence meaning “breast in partial involution with less than 50% glandular tissue”, combined with the feature “age” (*>* 50 vs *<* 50).

**Figure S10.**
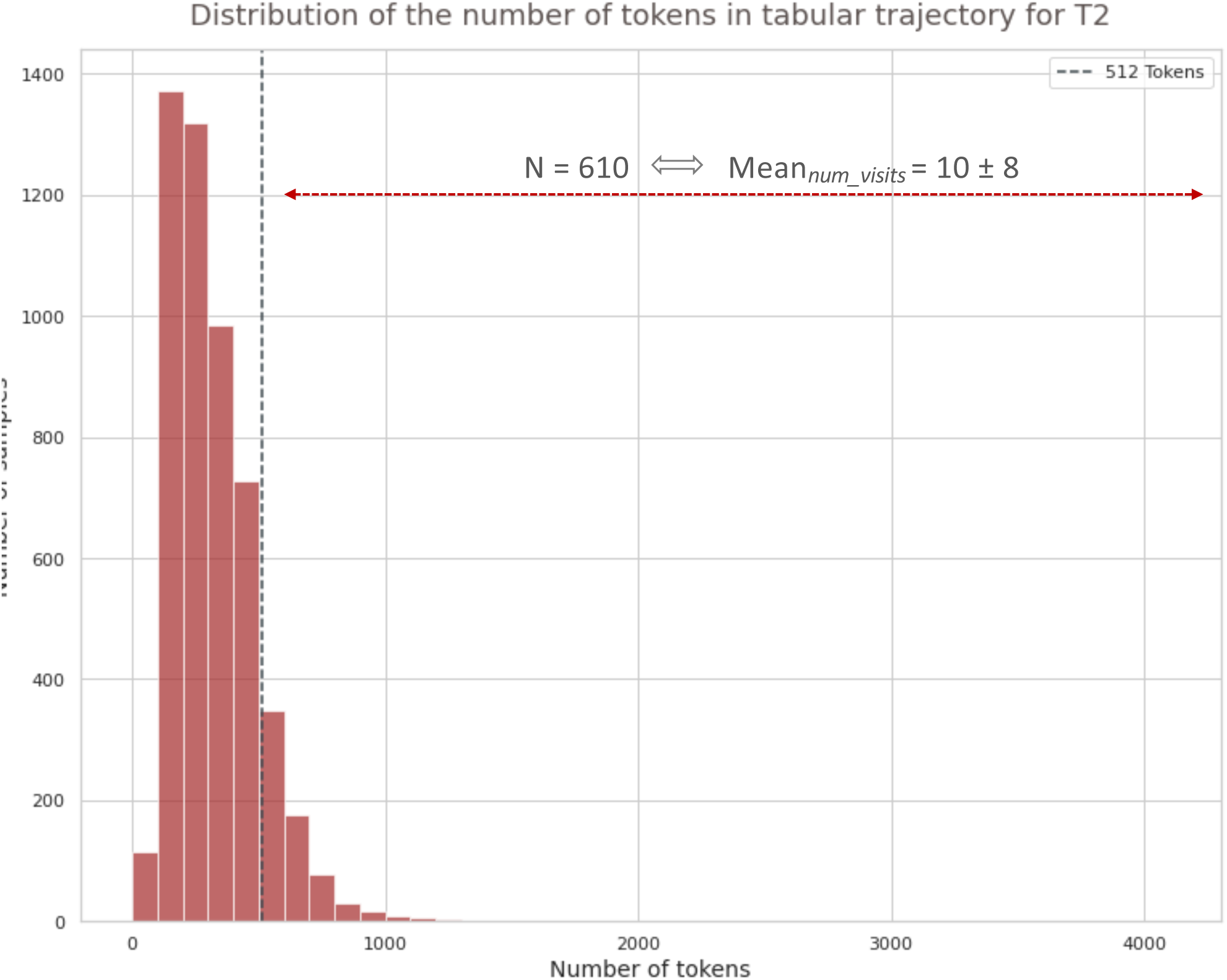
Distribution of the number of tokens per patient trajectory, for the prediction of disease-free survival 5 years after surgery. 610 samples exceed the maximum sequence length for Tabular BEHRT (512 tokens). This represents an average of 10 visits per patient that are not considered by Tabular BEHRT.

**Figure S11.**
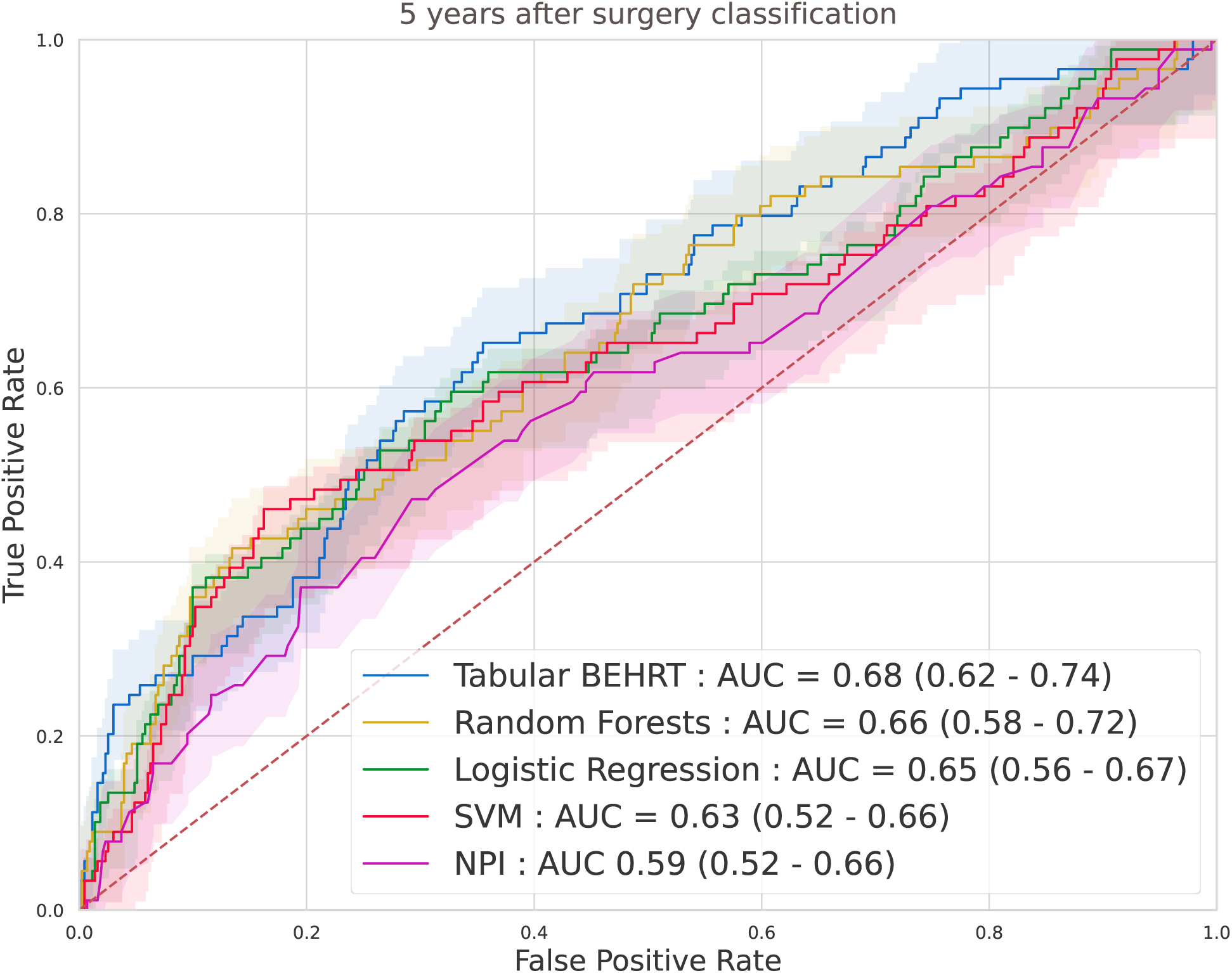
ROC curves for baselines and Tabular BEHRT, for predicting disease-free survival 5 years after surgery.

**Figure S12.**
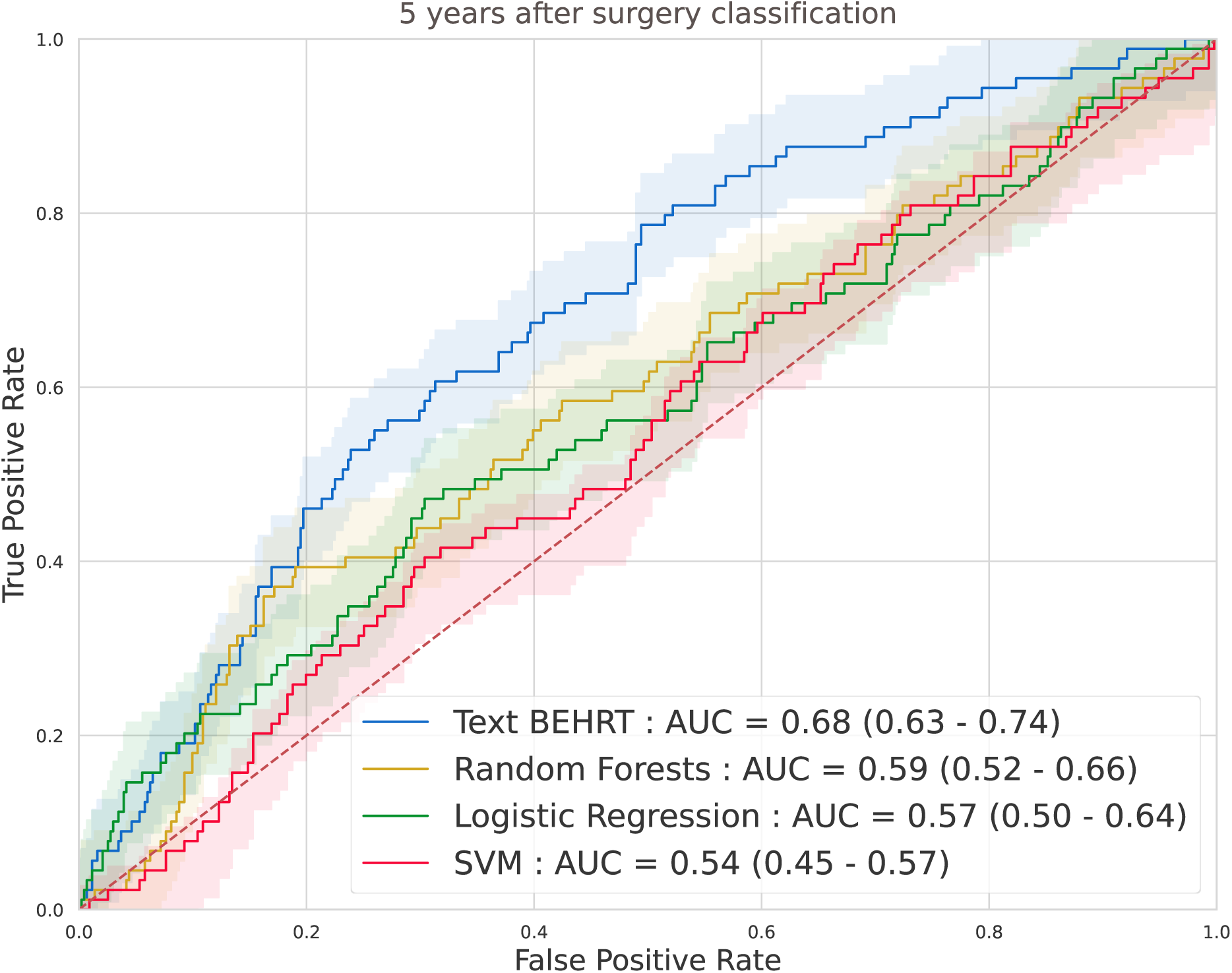
ROC curves for baselines and Text BEHRT, for predicting disease-free survival 5 years after surgery.

**Figure S13.**
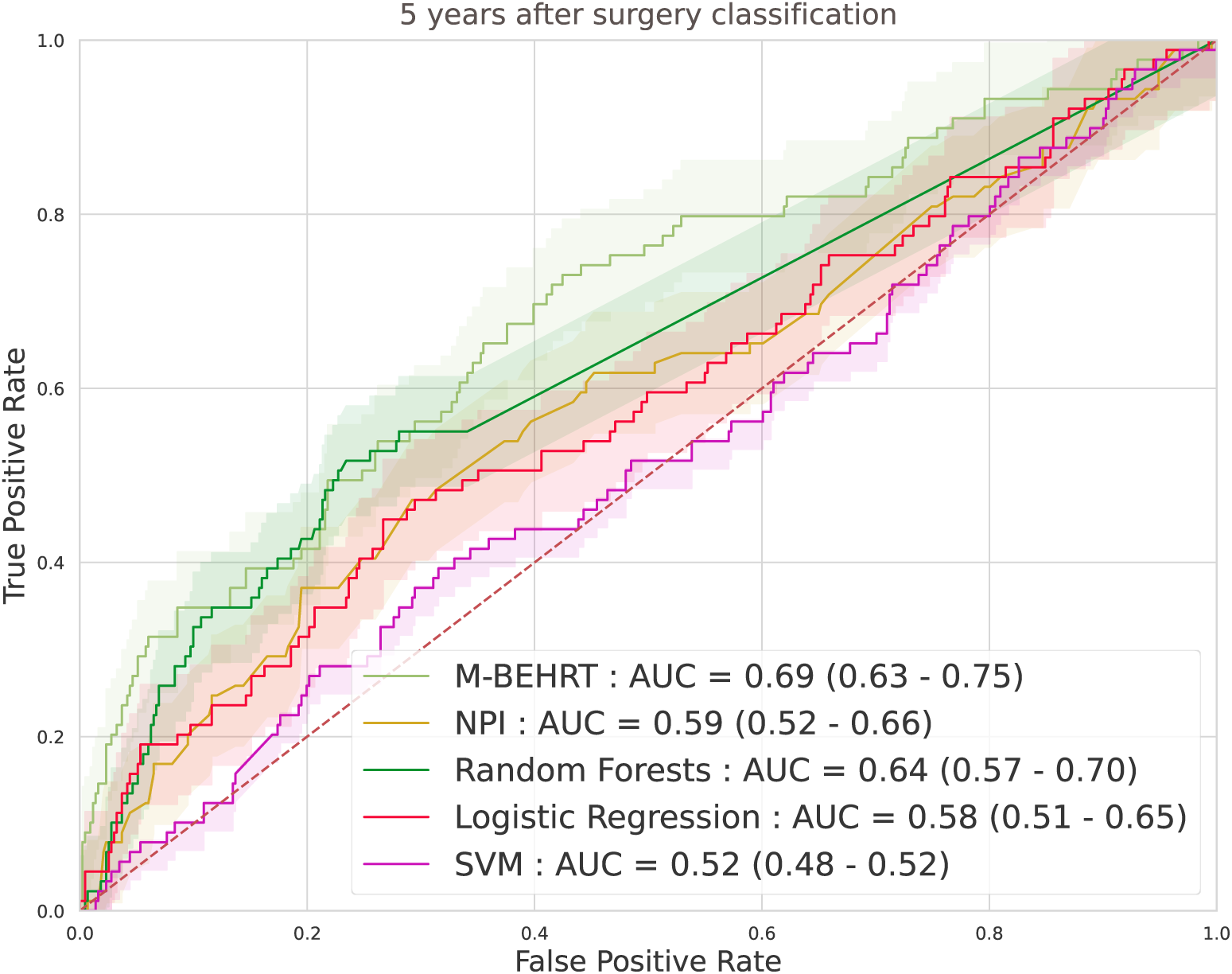
ROC curves M-BEHRT and baselines, for predicting disease-free survival 5 years after surgery.

**Figure S14.**
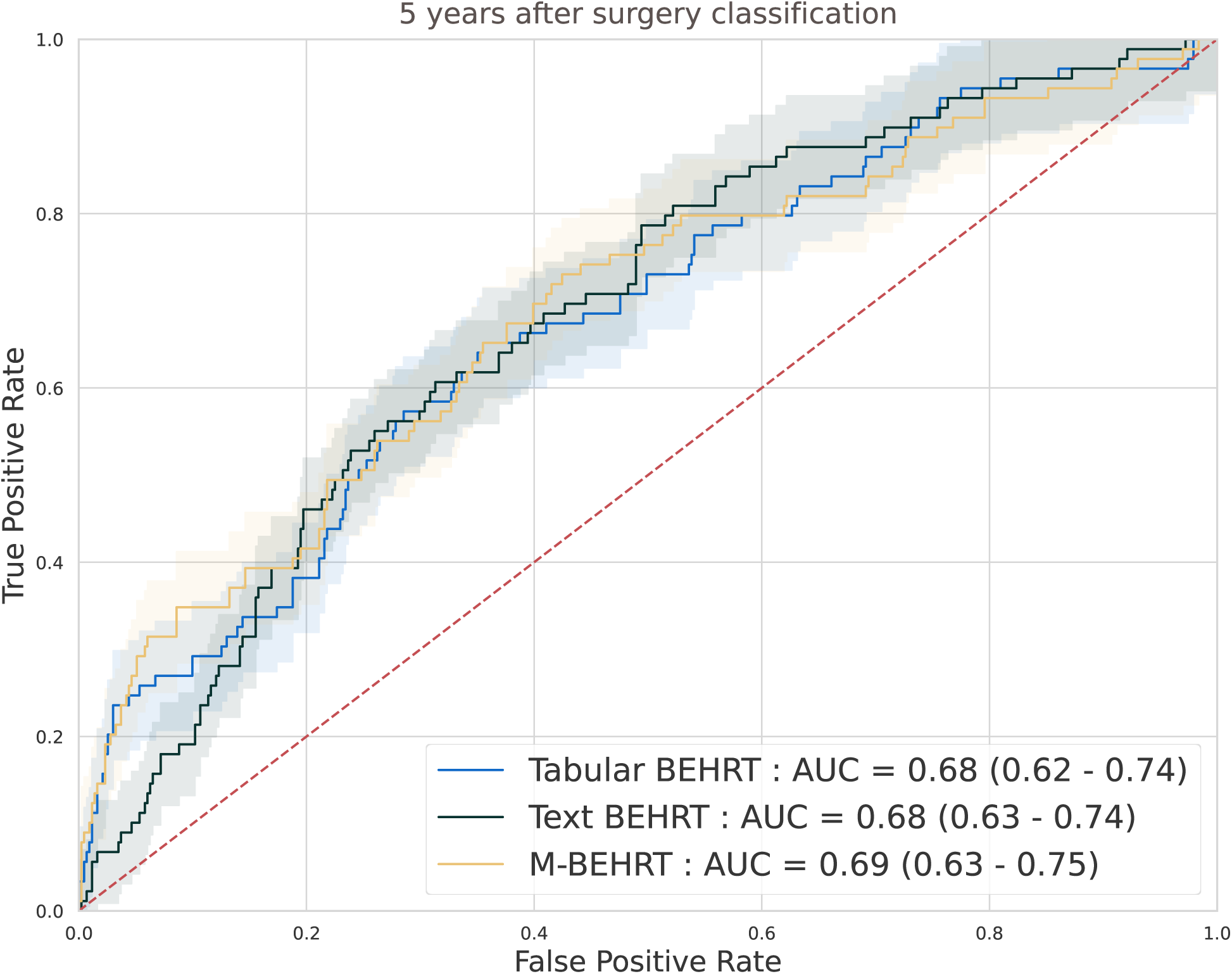
ROC curves comparing Tabular BEHRT and Text BEHRT against their combined model M-BEHRT, for the prediction of disease-free survival 5 years after surgery.

**Figure S15.**
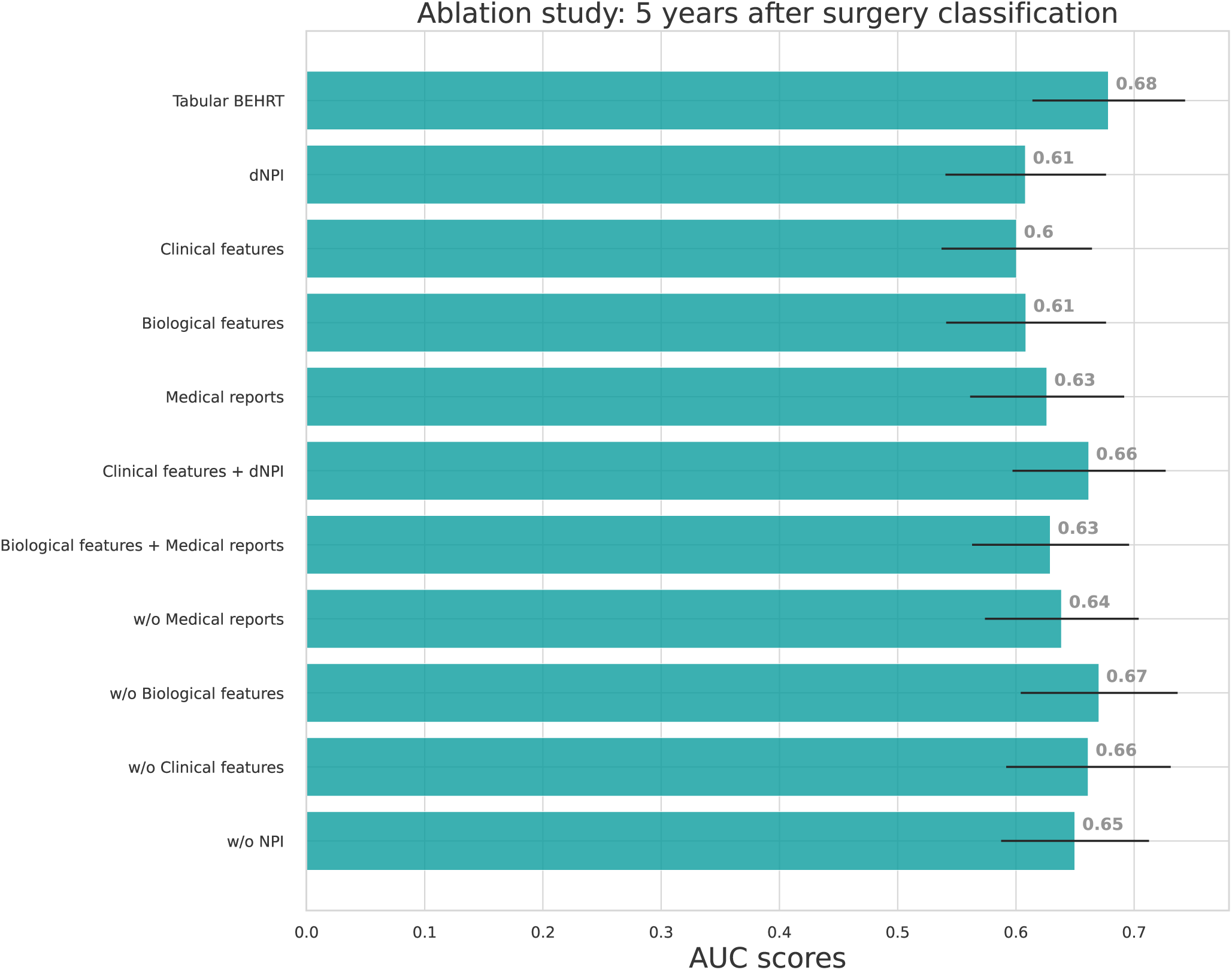
Ablation studies AUC-ROC on the test set for Tabular BEHRT, for the prediction of disease-free survival 5 years after surgery. We present results for the full model (Tabular BEHRT), then using only one of the 4 modalities (dNPI, clinical features, biological features, medical visits), two modalities (dNPI+clinical or biological+visits), then removing one of the 4 modalities. Here “medical records” stands for features extracted extracted from the medical record headers, that is to say, visit department and procedure. Performance scores are presented on the test set.

**Figure S16.**
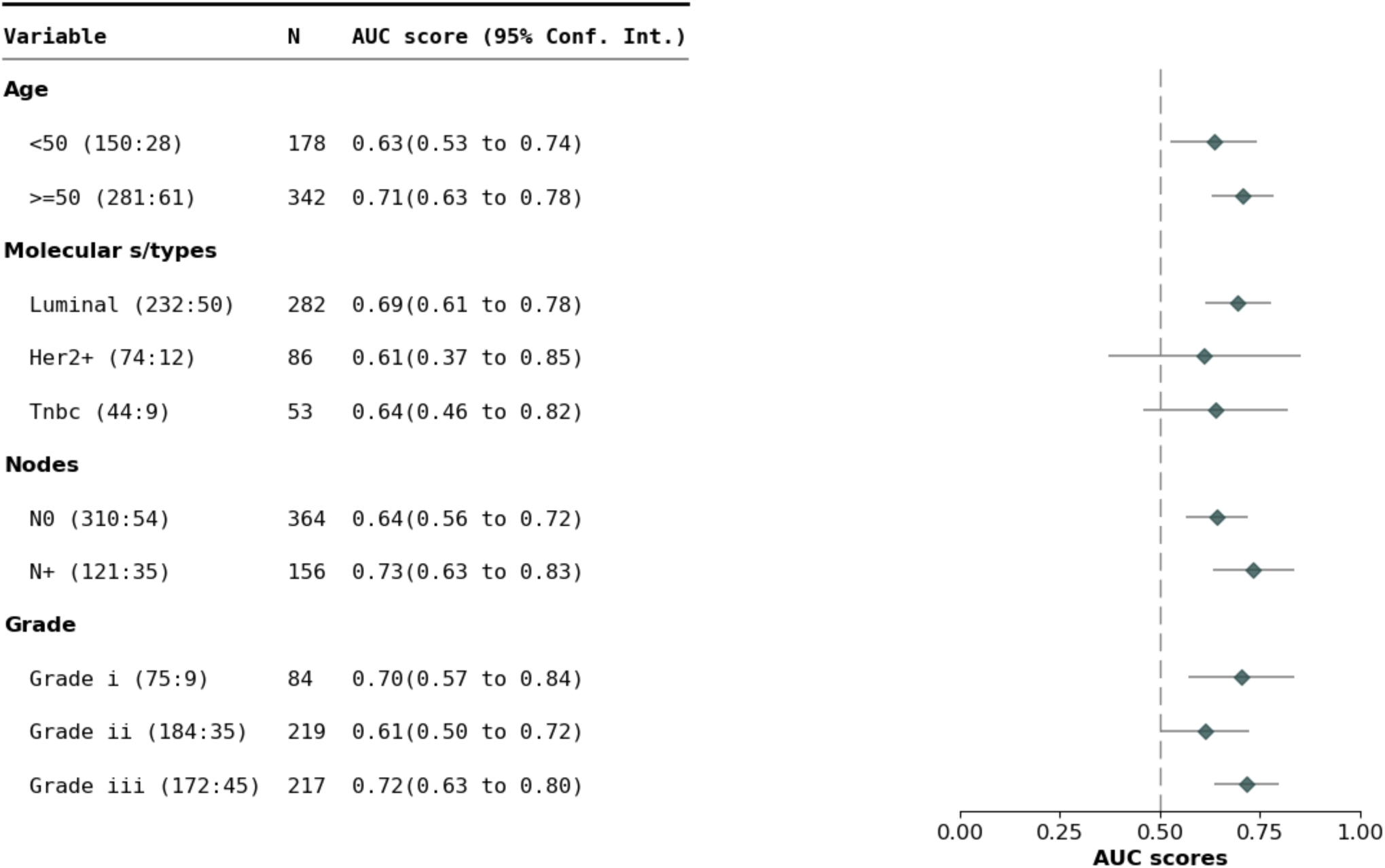
AUC-ROC of M-BEHRT stratified by patient age, cancer grade, molecular subtype and node status, for the prediction of disease-free survival 5 years after surgery.

**Figure S17.**
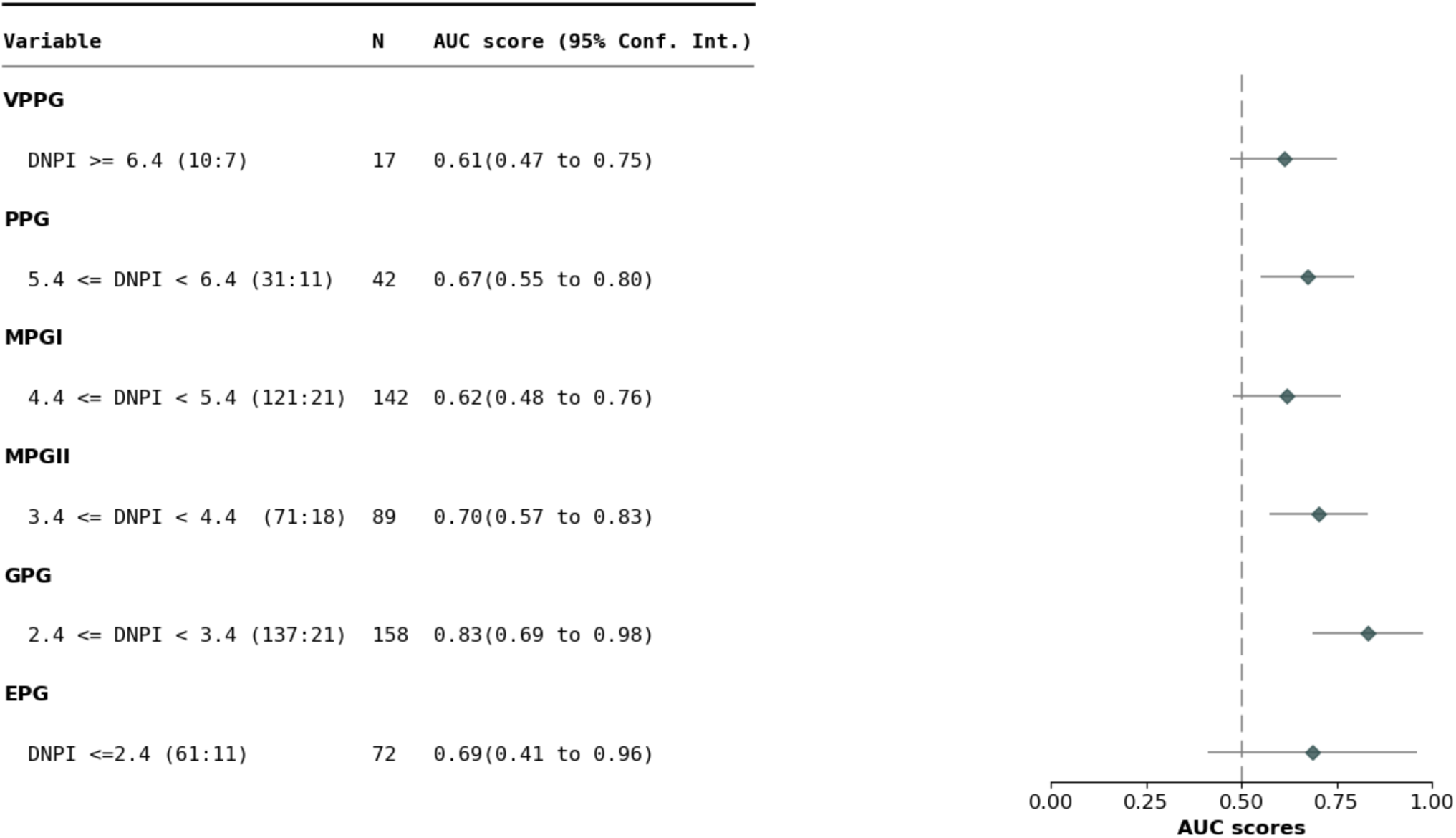
AUC-ROC of M-BEHRT stratified by NPI, for predicting disease-free survival 5 years after surgery.

**Figure S18.**
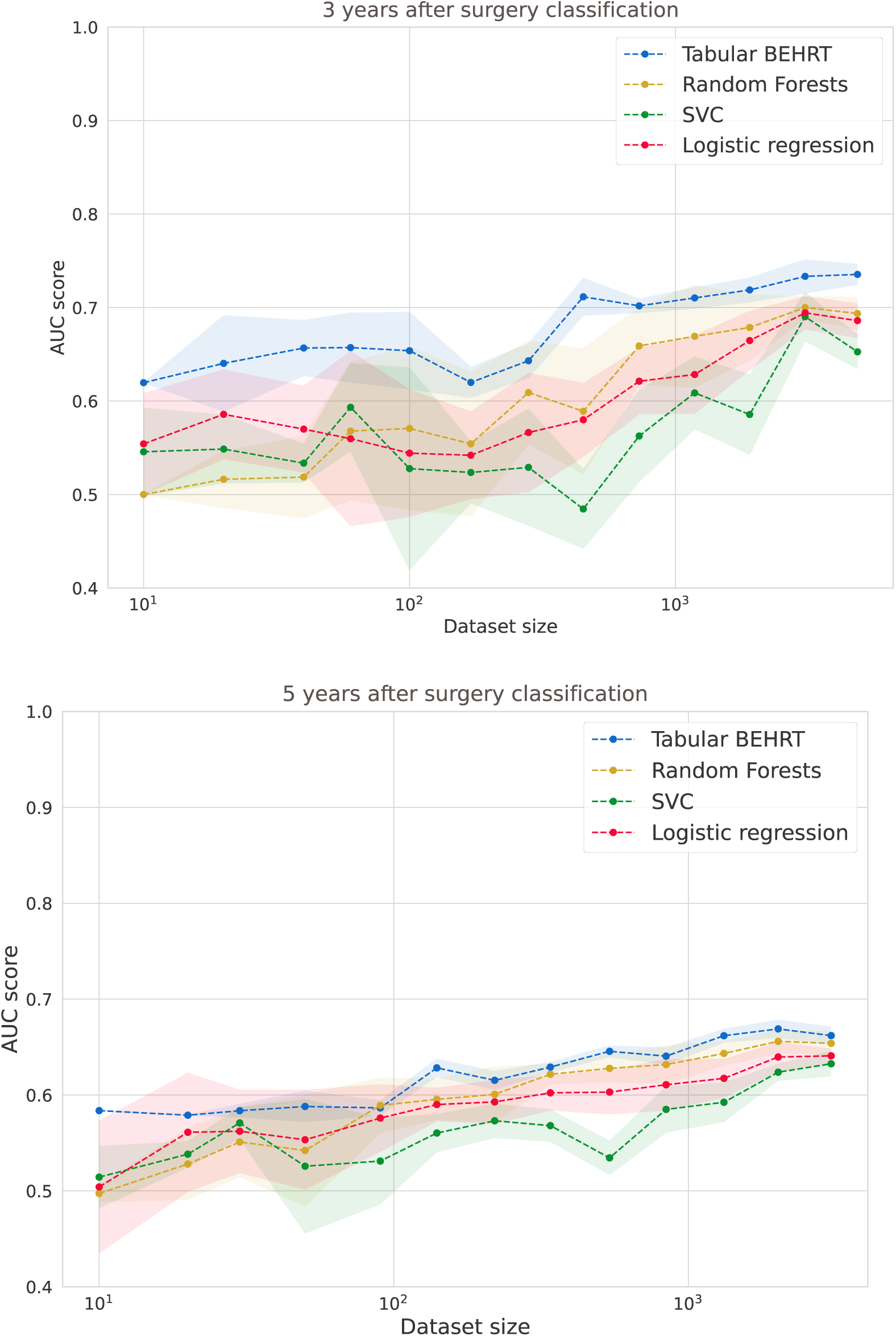
AUC-ROC on the test set of Tabular BEHRT, random forests, support vector classifier, and logistic regression trained on subsets of the training set of increasing sizes (x-axis), for the prediction of disease-free survival 3 (top) or 5 (bottom) years after surgery.

**Figure S19.**
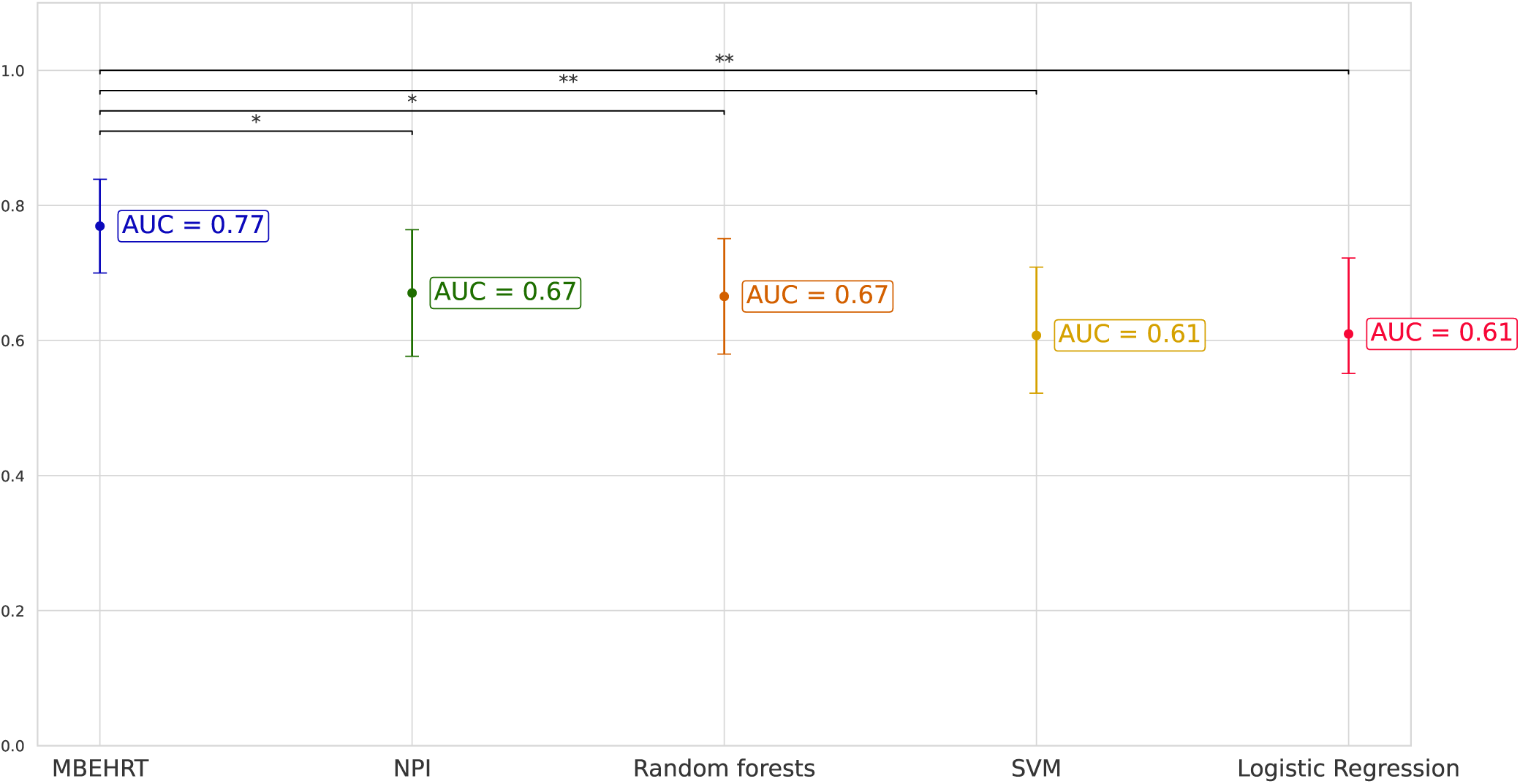
AUC scores comparison between M-BEHRT and the baselines for the prediction of disease-free survival 3 years after the surgery on the test set.

## Supplementary Text to Multimodal BEHRT: Transformers for Multimodal Electronical Health Records

### 1 PREPROCESSING OF FREE TEXT

Removing proper nouns is one of the key step of the preprocessing pipeline. This is important as specific doctor names may serve as proxy for the DFS classification, for example, when a doctor mostly handles severe cases. Patient names are already excluded from the reports, which had been anonymized before we accessed them. The first stage of this process consists in using part-of-speech tagging to remove proper nouns tags that follow titles such as *Dr*, *M.* (“Mr” in English), *Mme* (“Mrs” in English). However, proper nouns may appear without a title. We thus further constructed a list of proper nouns to remove from the text. We first built a list of names of Institut Curie’s health practicioners, obtained through the public directory of practicioners Cur (Accessed: 2023-01-30) as retrieved in 2023, and therefore only partially matching practicioners that were involved in the care of patients in the 2005–2012 period covered by our cohort). We additionally considered surnames given at least 30 times in France from 1891 to 2000 (n=218 912) and first names given at least 20 times from 1946 to 2022 in France (n=36 964), as provided by Institut National de La Statistique et des Etudes Econonomiques (INSEE) (Ins (Accessed: 2023-01-30), INS (Accessed: 2023-01-30)). We then removed from this list the proper names that correspond to disease names, such as Paget.

One other main difficulty that occur with free-text reports is the high number of typos. To address this issue, we used the pyspellchecker spell checking algorithm Barus (2023) which identifies, for each word of the corpus that is not found in a given dictionary, the most likely correct replacement for this presumably misspelled word. For effective spellchecking, it is crucial to have a rich dictionary that contains medical jargon. Therefore, we augmented the French vocabulary from OpenSubtitles Lison and Tiedemann (2016) (implemented by default in pyspellchecker) with the contents of the French open dictionary Usito ush (Accessed: 2023-01-30), as well as the 3 184 words from a French online medical dictionary Thomsen (Accessed: 2023-01-30), the CAS corpus of French clinical cases Grabar et al. (2018) which contains over 397 000 word occurences, a list of drug names in French vid (Accessed: 2023-01-30), and two lists of French medical abbreviations specific to oncology moz (2020); Poletto (2023). If, following this step, any words from the dictionary remain unidentified, we replaced them with the most likely correct spelling suggestion from Wikipedia wik (Accessed: 2023-01-30).

### 2 TEXT BEHRT INTERPRETATION

We choose to analyze the most frequent sequences for the DFS negative cohort that are not found in the DFS positive cohort. We ended up with the following sequences of words, some of which have been obtained with the overlapping resulting sequences:

- “sein en involution adipeuse partielle avec contingent glandulaire inferieur a 50”, (*breast in partial adipose involution with less than 50% glandular contingent*)
- “Traitement anterieur par hormone de croissance extractible non facteurs de risque de transmission de la mcj”, (*Previous treatment with extractable growth hormone without risk factors for mcj transmission*)
- “[avec] lymphadenectomie axillaire”, (*with axillary lymphadenectomy*)
- “syndrome de masse”, (*mass syndrom*)
- “[j1] solumedrol 80mg”, (*solumedrol 80mg*)
- “lovenox 0 4 ml”, (*lovenox 0 4 ml*)

